# Sugar-sweetened beverage and sugar intake during adolescence and risk of colorectal cancer precursors: a large prospective U.S. cohort study

**DOI:** 10.1101/2020.11.08.20227827

**Authors:** Hee-Kyung Joh, Dong Hoon Lee, Jinhee Hur, Katharina Nimptsch, Yoosoo Chang, Hyojee Joung, Xuehong Zhang, Leandro F. M. Rezende, Jung Eun Lee, Kimmie Ng, Yuan Chen, Jeffrey A. Meyerhardt, Andrew T Chan, Tobias Pischon, Mingyang Song, Charles S. Fuchs, Walter C. Willett, Yin Cao, Shuji Ogino, Edward Giovannucci, Kana Wu

## Abstract

**OBJECTIVE:** To examine the associations of adolescent sugar-sweetened beverage (SSB) and sugar intake with risk of colorectal cancer (CRC) precursors.

**DESIGN:** Prospective cohort study.

**SETTING:** Nurses’ Health Study II (1998-2015), United States.

**PARTICIPANTS:** 33106 women who completed a validated high school food frequency questionnaire about adolescent diet in 1998 and underwent lower gastrointestinal endoscopy between 1999 and 2015.

**MAIN OUTCOME MEASURES:** Incident CRC precursors confirmed by medical record review.

**RESULTS:** During follow-up, 2909 conventional adenoma, 1082 high-risk adenoma (≥1 cm in size, villous, high-grade dysplasia, or number ≥2), and 2355 serrated lesions were identified. Independent of adult intake, adolescent SSB and sugar intake was positively associated with risk of total and high-risk adenoma. Comparing ≥2 servings/day *v* <1 serving/week of SSB intake, multivariable odds ratios were 1.21 (95% confidence interval 1.00 to 1.47) for total and 1.21 (0.88 to 1.65) for high-risk adenoma. Per each 5% increment in calorie/day of total fructose intake, odds ratios were 1.17 (1.05 to 1.31) for total and 1.36 (1.14 to 1.62) for high-risk adenoma. By subsite, odds ratios were 1.25 (0.99 to 1.58) for proximal, 1.44 (1.12 to 1.84) for distal, and 1.74 (1.19 to 2.54) for rectal high-risk adenoma. Positive associations were stronger among women with low adolescent fruit, vegetable, or fiber intake. Among women with low fruit intake (<1.3 servings/day), odds ratios of total adenoma were 1.33 (1.11 to 1.59) for SSBs (≥1 serving/day *v* <1 serving/week) and 1.51 (1.26 to 1.82) for the highest quintile of total fructose (P≤0.024 for interaction). Neither SSB nor sugar intake during adolescence was associated with risk of serrated lesions.

**CONCLUSIONS:** Independent of adult intake, adolescent SSB and sugar intake was positively associated with colorectal adenoma, especially high-risk rectal adenoma. Our findings suggest that adolescence may be a critical developmental period of enhanced susceptibility to high sugar intake, possibly promoting precancerous lesions of CRC arising through the adenoma-carcinoma sequence.

**What is already known on this topic:**

- Relatively few studies have examined the association between sugar intake and colorectal neoplasia, and most prospective studies have reported null associations.
- Considering the long process of colorectal carcinogenesis and recent upward trends in early-onset colorectal cancer, early-life diet may be etiologically relevant.
- However, data on the relationship between high sugar intake during early-life and risk of colorectal neoplasia are lacking.

**What this study adds:**

- Higher intake of sugar-sweetened beverages (SSBs) and sugars during adolescence was significantly associated with increased risk of total and high-risk adenoma, especially high-risk rectal adenoma, but not serrated lesions.
- Positive associations were stronger among women with low fruit, vegetable, or fiber intake during adolescence.
- Our results suggest that limiting sugar intake and replacing SSBs with healthy alternatives during early-life may help to reduce risk of colorectal cancer precursors.

## INTRODUCTION

The global burden of colorectal cancer (CRC) is expected to increase to over 2.2 million new cases and 1.1 million cancer deaths per annum by 2030.^1^ The vast majority of CRC develops from malignant transformation of benign precursors, with the carcinogenesis process generally spanning several decades.^2 3^ CRC is among the most preventable cancers,^4^ with more than one-half of CRC cases and deaths attributable to modifiable diet and lifestyle factors.^5^ In several high-income countries, an emerging trend is the rising incidence of CRC at ages under 50 years, often referred to as early-onset CRC.^3 5 6^ In the U.S., incidence of early-onset CRC has been on the rise since the mid-1990s; and among those aged 50–64 years, CRC incidence has also started to increase since 2011 (after declines during the 2000s), reflecting elevated risk in generations born after 1950. This birth cohort effect indicates that shared population-level changes in exposures during early-life may have contributed to the recent increase in early-onset CRC.^5 6^

Sugar-sweetened beverages (SSBs) include carbonated and noncarbonated soft drinks, fruit drinks, and sports drinks, mostly sweetened with high-fructose corn syrup (first marketed in the early 1970s; usually 55% fructose and 45% glucose) or sucrose (table sugar; half fructose and half glucose).^7-9^ SSB intake is markedly increasing worldwide, particularly in developing countries,^10^ with concomitant increase in fructose and added sugar intake.^9^ In 53 low- and middle-income countries, 54% of adolescents aged 12–15 years consumed carbonated soft drinks at least once per day.^11^ In the U.S., SSBs were the largest source of added sugar, accounting for nearly a half of all added sugar intake in 2005–2006.^7^ One-half of adults and 63% of youths aged 2–19 years consumed at least 1 serving/day of SSBs, with the highest levels in adolescents aged 12–19 years, who consumed about 10% of daily calories from SSBs.^12 13^

SSB and high sugar intake can promote colorectal carcinogenesis by causing insulin resistance, obesity, and type 2 diabetes^14-18^—established risk factors for CRC.^4^ Despite the close link between insulin resistance and CRC,^19-22^ relatively few studies have examined the association between sugar intake and colorectal neoplasia, and most prospective studies have reported null associations.^4 23 24^ However, no previous study examined sugar intake during early-life. Considering the long process of carcinogenesis and recent upward trends in early-onset CRC,^3 6^ early-life diet may be etiologically relevant to CRC development.^25 26^ Moreover, adolescence is a unique growth period characterized by physiologically decreased insulin sensitivity and a surge of insulin-like growth factor 1 (IGF1).^27^ Thus, adolescence may be a critical developmental period of enhanced susceptibility to the adverse effects of high sugar intake. Our hypothesis was that high sugar intake during adolescence may play a role in early steps of colorectal carcinogenesis. We prospectively investigated the associations of adolescent SSB and sugar intake with risk of CRC precursors in a large cohort of young women.

## METHODS

### Study population

The Nurses’ Health Study II (NHSII) is an ongoing prospective cohort established in 1989 when 116430 U.S. female registered nurses aged 25–42 years returned a mailed questionnaire about various lifestyle factors and medical history.^28^ Follow-up questionnaires were mailed biennially to update the information and newly diagnosed diseases. In 1998, 45947 participants completed a high school food frequency questionnaire (HS-FFQ) about diet during adolescence.^29^ We included women who had completed the HS-FFQ in 1998 and subsequently underwent at least one lower gastrointestinal endoscopy between 1999 and 2015. We excluded women who had no lower bowel endoscopy during the follow-up because CRC precursors are generally asymptomatic and detected during an endoscopy. We also excluded women with a history of any cancer (other than nonmelanoma skin cancer), CRC precursors, Crohn’s disease, or ulcerative colitis prior to the return of the HS-FFQ, and those reporting implausible adolescent caloric intake (<600 or ≥5000 kcal/day), leaving a total of 33106 women for the current analyses. Then we additionally excluded individuals with missing values for each exposure variable of interest. The study protocol was approved by the institutional review boards of the Brigham and Women’s Hospital and Harvard T.H. Chan School of Public Health, and those of participating registries as required.

### Dietary assessment

Adolescent diet was assessed using a 124-item self-administered HS-FFQ, specifically designed to include food items commonly consumed between 1960 and 1982 when participants were 13– 18 years.^30^ Participants were asked how often, on average, they had consumed a standard portion size of each food or beverage when they were in high school, with 9 possible responses ranging from “never or less than once per month” to “6 or more times per day.” The reproducibility and validity of the HS-FFQ have been described previously.^29 31^ In brief, reproducibility at a 4-year interval was moderate-to-good (correlation for overall nutrients, *r*=0.65; foods, *r*=0.60; total fructose, *r*=0.65; cola, *r*=0.74; orange juice, *r*=0.74).^29^ Two independent validation studies reported adequate validity (*r* for overall nutrients, 0.40–0.58).^29 31^ Since 1991, adult diet was assessed every 4 years using a validated FFQ with approximately 131-food items.^32 33^

SSBs were defined as caffeinated and caffeine-free colas (e.g., Coke, Pepsi) and other carbonated (e.g., 7-Up) and non-carbonated sugary beverages (fruit punches, lemonades, or other fruit drinks). Artificially-sweetened beverages (ASBs) included carbonated and non-carbonated low-calorie or diet beverages. Standard serving sizes for SSBs and ASBs were 1 glass, a bottle, or a can (12 ounces). Fruit juice included orange, apple, grapefruit, and other fruit juices, with 1 small glass (6 ounces) as a serving size. Dairy products included milk, yogurt, cheese, ice cream, sherbet, milkshake, and frappe. Because disaccharide sucrose consists of half fructose and half glucose, total fructose intake was defined as the sum of free fructose and half of sucrose intake, and glucose intake from simple sugars was defined as the sum of free glucose and half of sucrose intake.^9^ Added sugar refers to sugar added to foods and beverages during processing or preparation.^34^ Total sugar represents the sum of monosaccharides and disaccharides (fructose, glucose, sucrose, and maltose).^34^

The nutrient database was primarily derived from U.S. Department of Agriculture sources,^29 35^ supplemented with information from manufacturers. Nutrient intake was adjusted for total energy intake using the residual method reported previously.^36-38^ For sugar intake, we also calculated nutrient density (percentage of daily calories contributed by each sugar).^38^ To better represent long-term diet during adulthood and reduce measurement error due to within-person variability,^37^ cumulative updated intake was calculated for adult diet by averaging the repeated measures from all available FFQs up to 2 years prior to the most recent endoscopy. As an indicator of overall diet during adolescence, we derived prudent and western dietary patterns using principal component analyses as reported previously.^39^ A western dietary pattern was characterized by high intake of desserts, sweets, snacks, red and processed meat, and refined grains; while a prudent pattern was characterized by high intake of vegetables, fruits, better-quality grains, fish, and poultry (Supplementary Table 1). For analyses of SSBs, ASBs, and fruit juice, dietary patterns were derived after excluding each beverage variable to avoid collinearity with the primary exposure.

### Outcome ascertainment

On each biennial questionnaire, participants were asked whether they underwent a lower bowel endoscopy and the reasons why (screening, family history of CRC, symptoms), and whether CRC or precursors were diagnosed. Participants who reported a diagnosis of CRC precursors were asked for permission to access medical and pathological records. Physicians blinded to participant exposure information reviewed the records to verify the diagnosis and accrue information on CRC precursor size, number, subtype (conventional adenoma [hereinafter referred to as adenoma], serrated lesion), subsite (proximal, distal, rectal), and histology (tubular, tubulovillous, villous; with or without high-grade dysplasia). We subdivided adenoma into high-risk (≥1 cm, any villous histology, high-grade dysplasia, or ≥2 adenomas) and low-risk (<1 cm, tubular, and single) adenoma.^40^ Serrated lesions included hyperplastic polyps, sessile serrated adenoma/sessile serrated polyp, and traditional serrated adenoma,^41^ with subdivision by size (<1cm, ≥1 cm).

### Statistical analysis

SSB and other beverage intake was grouped into 4 categories: <1 serving/week, 1–6 servings/week, 1 serving/day, and ≥2 servings/day. Sugar intake was categorized into quintiles using either nutrient density or energy-adjusted intake. SSB and sugar intake was also treated as continuous variables. Time-varying covariates were updated to 2 years prior to most recent endoscopy, whenever available. To handle individuals with multiple endoscopies and time-varying covariates efficiently, the Andersen-Gill data structure was used.^42^ A new dataset were generated for each questionnaire cycle when participants reported an endoscopy. Thus, participants with multiple endoscopies during follow-up could provide multiple records. Once CRC precursor(s) were diagnosed, the participant was censored.

Odds ratios and 95% confidence intervals of CRC precursors were estimated using logistic regression for clustered data (SAS PROC GENMOD) where each participant represented a cluster. We constructed 3 multivariable models with adjustment for various potential confounders during both adolescence and adulthood.^4 43^ Model 1 included age, time period of endoscopy, time since most recent endoscopy, number of endoscopies, and reason for endoscopy. Model 2 was additionally adjusted for family history of CRC in first degree of relatives, menopausal status/menopausal hormone use, current aspirin use ≥2 times/week, history of type 2 diabetes, adult height, body mass index (BMI, at 18 years and current), smoking status (adolescent and current), alcohol consumption (18–22 years and current), and physical activity (adolescent and current). In Model 3, to assess whether associations were independent of other dietary factors and overall unhealthy dietary pattern, we further adjusted for adolescent and adult dietary intake (total calorie, calcium, vitamin D, folate, fiber, fruits, vegetables, and dairy), current total red meat intake, a western dietary pattern score during adolescence, and corresponding adult variables to adolescent exposure variables.

Tests for trend were performed by assigning a median value to each category of exposure variables and modeling this value as a continuous variable, with the Wald test used to assess statistical significance. Stratified analyses were performed to examine whether associations varied across strata of known CRC risk factors during adolescence (e.g., family history of CRC, BMI, physical activity, fruit and vegetable intake).^43^ Tests for interaction were performed by including cross-product terms of exposure and stratification variables in the model and utilizing a Wald test. To compare the effects of diet during different life stages, we examined joint associations of adolescent and adult intake with CRC precursor risk. The effects of substituting fruits, fruit juice, or dairy for SSBs were estimated by simultaneously including both SSBs and one of these food items as continuous variables in models; ORs and 95% CIs were calculated from the differences in coefficients and corresponding variances and covariances.^44-46^ All tests were two-sided with P<0.05 considered statistically significant. All analyses were performed using SAS version 9.4 (SAS Institute, Cary, NC).

### Patient and public involvement

No patients were involved in setting the research question or the outcome measures, nor were they involved in the design and implementation of the study. We plan to disseminate these findings to participants through the study websites (www.nurseshealthstudy.org) and annual newsletter and to the general public in a press release.

## RESULTS

Baseline characteristics of the participants are described in table 1. The mean age of the participants was 44.1±4.5 years when the HS-FFQ was completed. During adolescence, 12.6% of women had consumed ≥1 serving/day of SSBs (≥2 servings/day, 4.8%), while in adulthood, 11.1% consumed ≥1 serving/day (≥2 servings/day, 3.2%). Adolescent SSB intake contributed to 2.6% of daily calories. SSB-source added sugar intake was 18.2 g/day, on average, which was lower than the mean intake (27 g/day) among U.S. youths aged 1–18 years in the national survey in 1971– 1974,^47^ the median time period when our participants attended high school. Participants with higher adolescent SSB and total fructose intake were more likely to be physically active during adolescence. While women with higher adolescent SSB intake were more likely to have higher red meat intake and lower fruit and vegetable intake, those with higher fructose intake were more likely to have lower red meat and higher fruit and vegetable intake during both adolescence and adulthood. The correlation between SSB and total fructose intake during adolescence was low-to-modest (Spearman correlation, *r*=0.38; supplementary table 2). Adolescent diet was only weakly correlated with adult diet (SSBs, age-corrected partial *r*=0.25; total fructose, *r*=0.27).

**Table 1.**
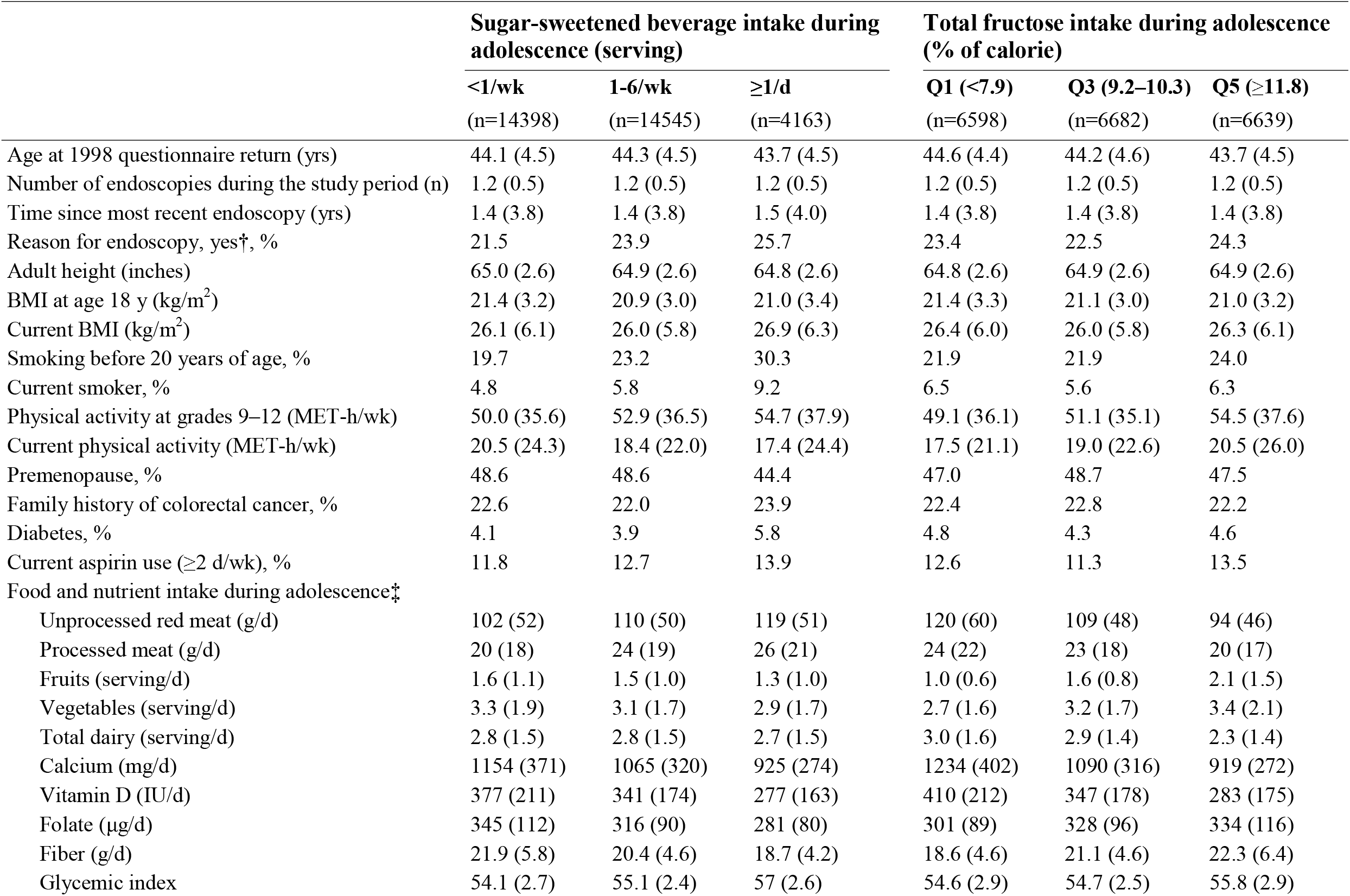
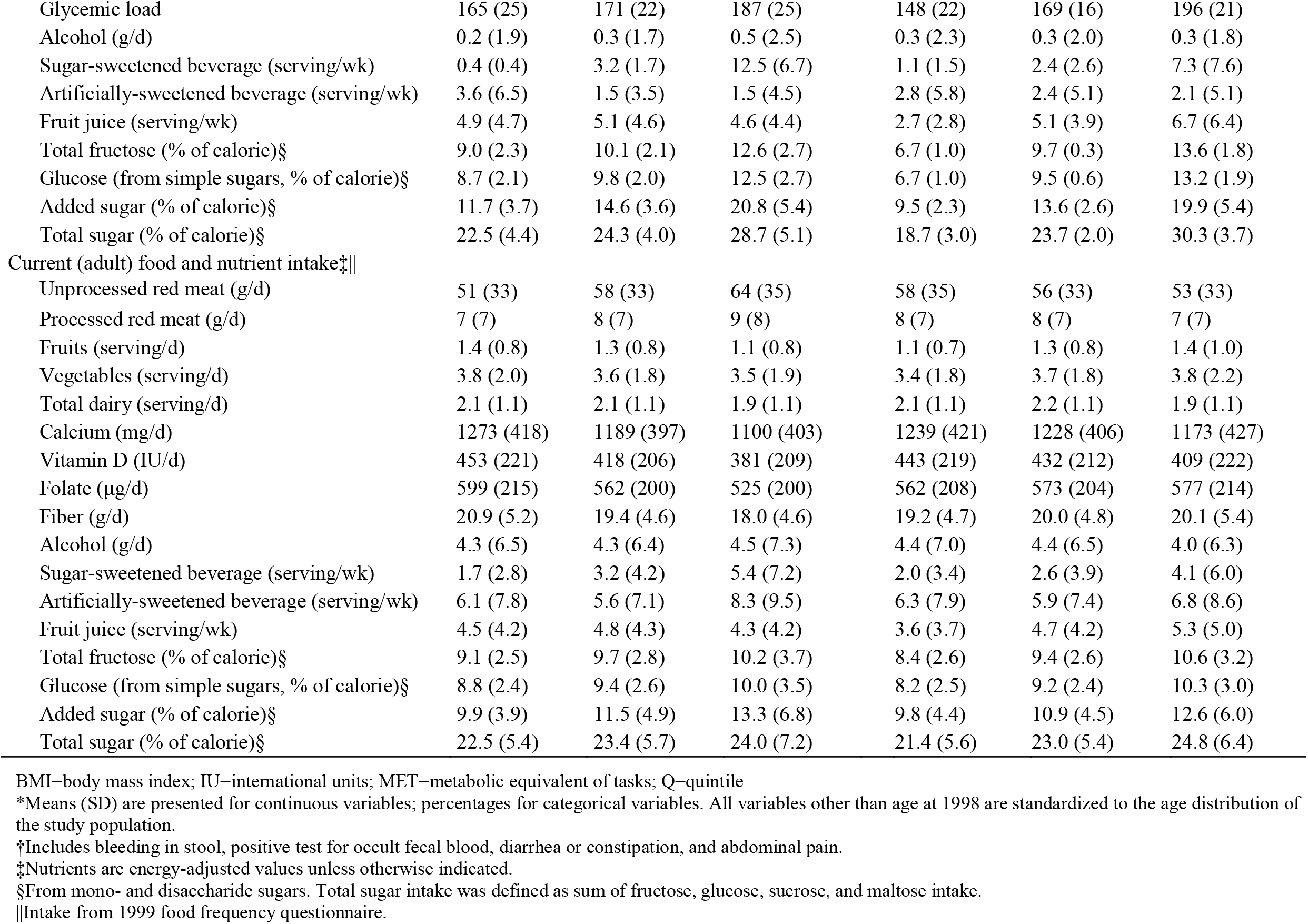
**Baseline characteristics of participants by sugar-sweetened beverage and total fructose intake during adolescence in the Nurses’ Health Study II\***

During follow-up, 4744 women were diagnosed with at least one CRC precursor, of whom 2909 had at least one adenoma (1548 proximal, 1205 distal, 458 rectal, and 1082 high-risk adenoma), and 2355 at least one serrated lesion. The mean age at diagnoses was 52.2±4.3 years (range, 37–68).

### SSB and sugar intake and CRC precursor risk

Independent of adult intake, higher intake of SSBs and total fructose during adolescence was significantly associated with increased risk of total adenoma (tables 2 and 3). For fructose intake, positive associations with adenoma risk were not significant in model 1 and 2. However, additional adjustment for dietary covariables (especially adolescent fruit, fiber, and calcium intake) in model 3 substantially stregthened the associations. In fully adjusted models, the odd ratios of total adenoma were 1.21 (95% confidence interval 1.00 to 1.47; P=0.012 for trend) for SSB intake, comparing ≥2 servings/day *v* <1 serving/week, and 1.17 (1.05 to 1.31; P=0.006 for trend) per 5% of calorie/day increase in total fructose intake. By subsite, each serving per day increment in SSBs was associated with higher risk of proximal (odds ratio 1.13, 95% confidence interval 1.02 to 1.26), distal (1.08, 0.95 to 1.21), and rectal (1.28, 1.07 to 1.53) adenoma; a 5% calorie/day increment in total fructose was positively associated with proximal (1.12, 0.96 to 1.30), distal (1.24, 1.05 to 1.48) and rectal (1.43, 1.10 to 1.86) adenoma. Results for glucose (from simple sugars), added sugar, and total sugar were similar to the results for total fructose, but effect sizes were slightly smaller than for total fructose (supplementary table 3). Neither SSB nor sugar intake during adolescence was associated with risk of total and large serrated lesions (all P≥0.34 for trend). We found no significant associations between adolescent ASB or fruit juice intake and CRC precursor risk (supplementary table 4). Contrary to adolescent intake, SSB and sugar intake during adulthood was not associated with adenoma risk. For adult SSB intake (≥2 servings/day *v* <1 serving/week), odd ratios were 0.87 (95% confidence interval 0.67 to 1.14) for total and 0.87 (0.47 to 1.61) for rectal adenoma; for adult total fructose intake (highest *v* lowest quintile), odd ratios were 0.95 (0.82 to 1.09) for total and 0.79 (0.56 to 1.12) for rectal adenoma.

**Table 2.**
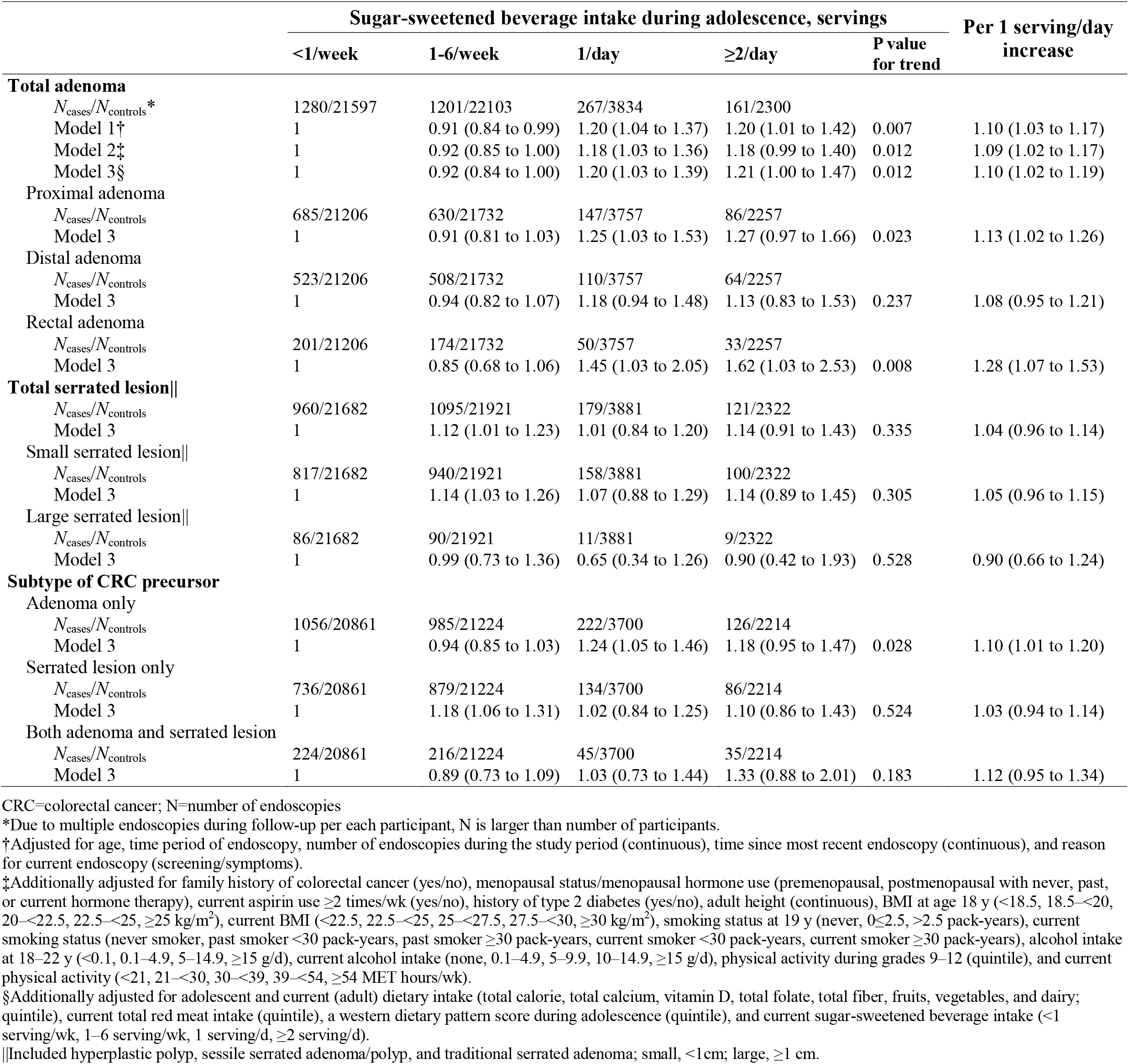
**Odds ratios and 95% confidence intervals of CRC precursor according to sugar-sweetened beverage intake during adolescence in the Nurses’ Health Study II, 1998-2015**

**Table 3.** **Odds ratios and 95% confidence intervals of CRC precursor according to total fructose intake during adolescence in the Nurses’ Health Study II, 1998–2015**

|  | Total fructose intake during adolescence, % of calorie |  |  |  |  |  | Per 5% of calorie/day increase |
| --- | --- | --- | --- | --- | --- | --- | --- |
| | Q1<br>($<7.9$) | Q2<br>($7.9\text{--}9.2$) | Q3<br>($9.2\text{--}10.3$) | Q4<br>($10.3\text{--}11.8$) | Q5<br>($\geq 11.8$) | P value<br>for trend | |
| **Total adenoma** |  |  |  |  |  |  |  |
| $N_{\text{cases}}/N_{\text{controls}}$ * | 573/9976 | 570/9978 | 595/9954 | 591/9958 | 580/9968 | | |
| Model 1† | 1 | 1.00 (0.89 to 1.12) | 1.05 (0.93 to 1.18) | 1.05 (0.93 to 1.18) | 1.03 (0.92 to 1.16) | 0.436 | 1.04 (0.95 to 1.13) |
| Model 2‡ | 1 | 1.00 (0.89 to 1.13) | 1.06 (0.94 to 1.20) | 1.07 (0.95 to 1.20) | 1.06 (0.94 to 1.20) | 0.207 | 1.06 (0.97 to 1.16) |
| Model 3§ | 1 | 1.05 (0.93 to 1.19) | 1.14 (1.00 to 1.30) | 1.19 (1.04 to 1.36) | 1.20 (1.04 to 1.39) | 0.006 | 1.17 (1.05 to 1.31) |
| Proximal adenoma |  |  |  |  |  |  |  |
| $N_{\text{cases}}/N_{\text{controls}}$ | 316/9795 | 306/9807 | 323/9765 | 310/9776 | 293/9809 | | |
| Model 3 | 1 | 1.03 (0.87 to 1.22) | 1.15 (0.96 to 1.36) | 1.15 (0.96 to 1.39) | 1.13 (0.92 to 1.38) | 0.164 | 1.12 (0.96 to 1.30) |
| Distal adenoma |  |  |  |  |  |  |  |
| $N_{\text{cases}}/N_{\text{controls}}$ | 239/9795 | 221/9807 | 241/9765 | 251/9776 | 253/9809 | | |
| Model 3 | 1 | 0.97 (0.80 to 1.18) | 1.10 (0.90 to 1.35) | 1.21 (0.98 to 1.48) | 1.25 (1.00 to 1.56) | 0.014 | 1.24 (1.05 to 1.47) |
| Rectal adenoma |  |  |  |  |  |  |  |
| $N_{\text{cases}}/N_{\text{controls}}$ | 77/9795 | 97/9807 | 84/9765 | 101/9776 | 99/9809 | | |
| Model 3 | 1 | 1.39 (1.01 to 1.89) | 1.24 (0.90 to 1.73) | 1.61 (1.15 to 2.25) | 1.62 (1.14 to 2.31) | 0.008 | 1.43 (1.10 to 1.86) |
| **Total serrated lesion\|\|** |  |  |  |  |  |  |  |
| $N_{\text{cases}}/N_{\text{controls}}$ | 479/9951 | 442/9993 | 486/9945 | 486/9934 | 462/9983 | | |
| Model 3 | 1 | 0.93 (0.80 to 1.07) | 1.03 (0.89 to 1.20) | 1.06 (0.91 to 1.23) | 1.01 (0.86 to 1.19) | 0.486 | 1.04 (0.92 to 1.18) |
| Small serrated lesion\|\| |  |  |  |  |  |  |  |
| $N_{\text{cases}}/N_{\text{controls}}$ | 405/9951 | 374/9993 | 420/9945 | 430/9934 | 386/9983 | | |
| Model 3 | 1 | 0.93 (0.80 to 1.08) | 1.06 (0.91 to 1.24) | 1.12 (0.96 to 1.32) | 1.01 (0.85 to 1.21) | 0.408 | 1.06 (0.93 to 1.21) |
| Large serrated lesion\|\| |  |  |  |  |  |  |  |
| $N_{\text{cases}}/N_{\text{controls}}$ | 46/9951 | 41/9993 | 40/9945 | 29/9934 | 40/9983 | | |
| Model 3 | 1 | 0.88 (0.57 to 1.37) | 0.85 (0.54 to 1.35) | 0.60 (0.36 to 1.00) | 0.86 (0.54 to 1.37) | 0.345 | 0.83 (0.56 to 1.22) |
| **Subtype of CRC precursor** |  |  |  |  |  |  |  |
| Adenoma only |  |  |  |  |  |  |  |
| $N_{\text{cases}}/N_{\text{controls}}$ | 472/9598 | 488/9618 | 482/9581 | 469/9594 | 478/9608 | | |
| Model 3 | 1 | 1.10 (0.96 to 1.26) | 1.13 (0.98 to 1.30) | 1.15 (0.99 to 1.34) | 1.20 (1.03 to 1.41) | 0.027 | 1.15 (1.02 to 1.30) |
| Serrated lesion only |  |  |  |  |  |  |  |
| $N_{\text{cases}}/N_{\text{controls}}$ | 378/9598 | 360/9618 | 373/9581 | 364/9594 | 360/9608 | | |
| Model 3 | 1 | 0.96 (0.82 to 1.12) | 1.00 (0.85 to 1.17) | 0.99 (0.84 to 1.17) | 0.99 (0.83 to 1.18) | 0.998 | 1.00 (0.87 to 1.15) |
| Both adenoma and serrated lesion |  |  |  |  |  |  |  |
| $N_{\text{cases}}/N_{\text{controls}}$ | 101/9598 | 82/9618 | 113/9581 | 122/9594 | 102/9608 | | |
| Model 3 | 1 | 0.84 (0.62 to 1.14) | 1.20 (0.89 to 1.61) | 1.36 (1.01 to 1.85) | 1.20 (0.85 to 1.67) | 0.061 | 1.28 (0.99 to 1.65) |

CRC=colorectal cancer; N=number of endoscopies; Q=quintile
\*Due to multiple endoscopies during follow-up per each participant, N is larger than number of participants.
†Adjusted for age, time period of endoscopy, number of endoscopies during the study period (continuous), time since most recent endoscopy (continuous), and reason for current endoscopy (screening/symptoms).
‡Additionally adjusted for family history of colorectal cancer, menopausal status/menopausal hormone use, current aspirin use $\geq 2$ times/wk, history of type 2 diabetes, adult height (continuous), BMI at age 18 y, current BMI, smoking status at 19 y, current smoking status, alcohol intake at 18–22 y, current alcohol intake, physical activity during grades 9–12, and current physical activity.
§Additionally adjusted for adolescent and current dietary intake (total calorie, total calcium, vitamin D, total folate, total fiber, fruits, vegetables, and dairy), current total red meat intake, a western dietary pattern score during adolescence, and current total fructose intake.
||Included hyperplastic polyp, sessile serrated adenoma/polyp, and traditional serrated adenoma; small, <1cm; large, $\geq 1$ cm.

### SSB and sugar intake and risk of high-risk adenoma

Adolescent intake of SSBs and total fructose was positively associated with high-risk adenoma, but not with low-risk adenoma (table 4). The multivariable odds ratios of high-risk adenoma were 1.21 (95% confidence interval 0.88 to 1.65; P=0.085 for trend) for SSB intake, comparing ≥2 servings/day *v* <1 serving/week, and 1.36 (1.14 to 1.62; P=0.001 for trend) per each 5% increase in calorie/day of total fructose. By subsite, associations of SSB intake were significant in the rectum (per 1 serving/day, 1.48, 1.16 to 1.88; P=0.001 for trend), but not in the proximal and distal colon (P≥0.157 for trend). Per 5% increment in calorie/day, fructose intake was positively associated with high-risk proximal (1.25, 0.99 to 1.58), distal (1.44, 1.12 to 1.84) and rectal (1.74, 1.19 to 2.54) adenoma. Results for glucose (from simple sugars), added sugar, and total sugar were similar (but somewhat weaker) to the results for total fructose (supplementary table 3). Adult sugar intake was not associated with high-risk adenoma with odds ratios of 0.92 (95% confidence interval 0.61 to 1.38) for SSBs (≥2 servings/day *v* <1 serving/week) and 0.93 (0.75 to 1.16) for total fructose (highest *v* lowest quintile).

**Table 4.**
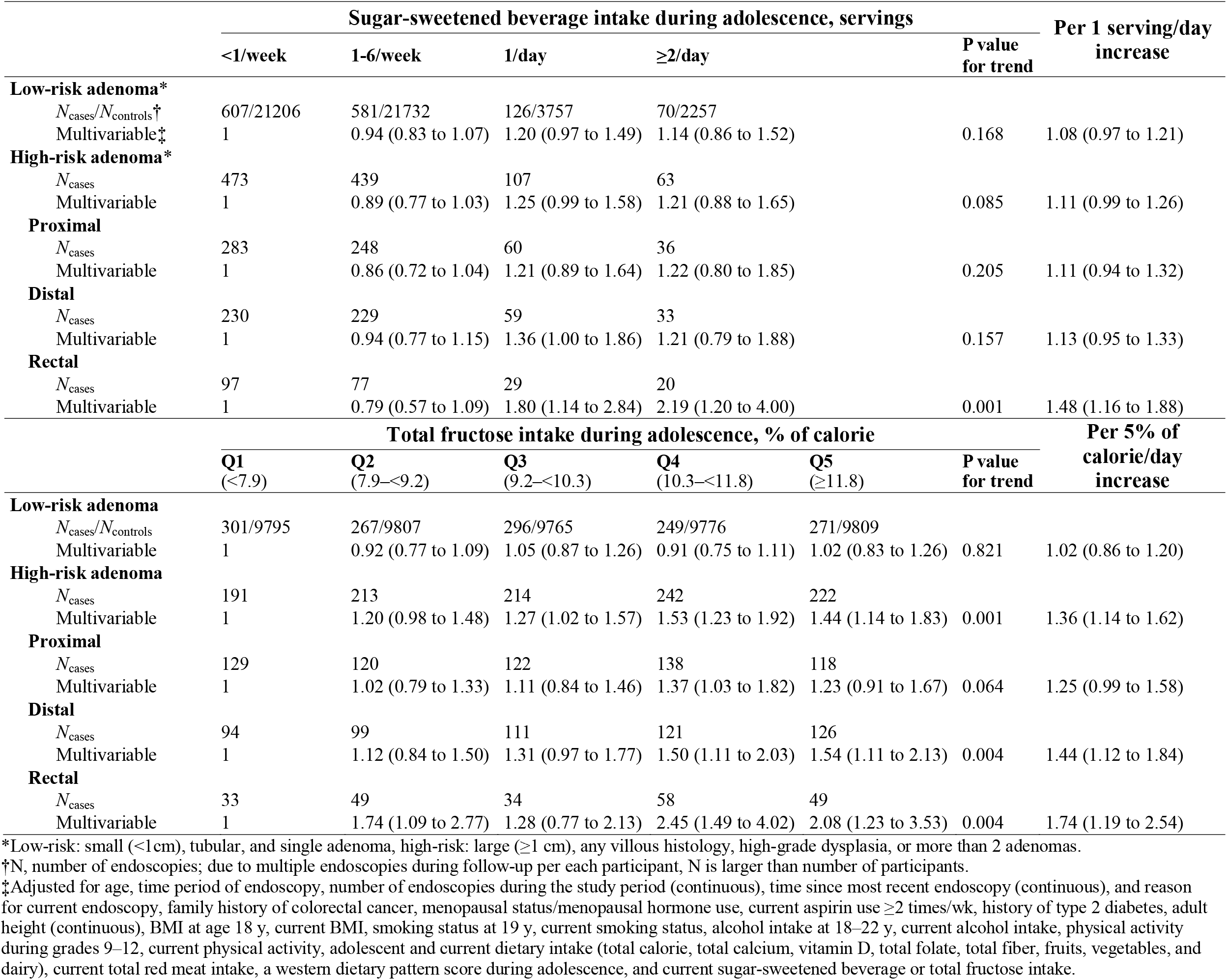
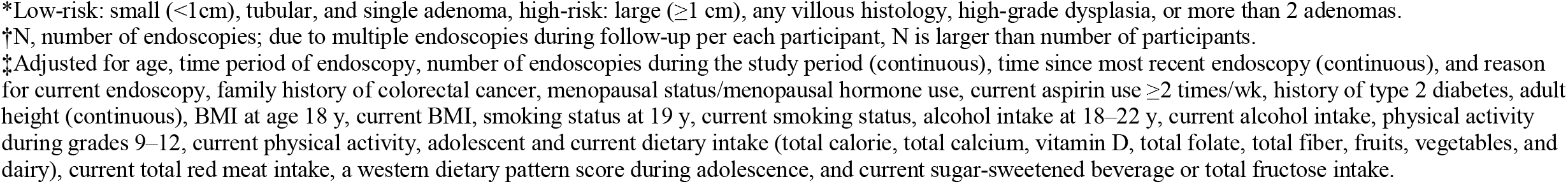
**Multivariable odds ratios and 95% confidence intervals of low- and high-risk colorectal adenoma according to sugar-sweetened beverage and total fructose intake during adolescence in the Nurses’ Health Study II, 1998-2015**

|  | Sugar-sweetened beverage intake during adolescence, servings |  |  |  |  | P value<br>for trend | Per 1 serving/day<br>increase |
| --- | --- | --- | --- | --- | --- | --- | --- |
|  | <1/week | 1-6/week | 1/day | ≥2/day |  |  |  |
| **Low-risk adenoma\*** |  |  |  |  |  |  |  |
| *N*<sub>cases</sub>/*N*<sub>controls</sub><sup>†</sup> | 607/21206 | 581/21732 | 126/3757 | 70/2257 |  |  |  |
| Multivariable<sup>‡</sup> | 1 | 0.94 (0.83 to 1.07) | 1.20 (0.97 to 1.49) | 1.14 (0.86 to 1.52) |  | 0.168 | 1.08 (0.97 to 1.21) |
| **High-risk adenoma\*** |  |  |  |  |  |  |  |
| *N*<sub>cases</sub> | 473 | 439 | 107 | 63 |  |  |  |
| Multivariable | 1 | 0.89 (0.77 to 1.03) | 1.25 (0.99 to 1.58) | 1.21 (0.88 to 1.65) |  | 0.085 | 1.11 (0.99 to 1.26) |
| **Proximal** |  |  |  |  |  |  |  |
| *N*<sub>cases</sub> | 283 | 248 | 60 | 36 |  |  |  |
| Multivariable | 1 | 0.86 (0.72 to 1.04) | 1.21 (0.89 to 1.64) | 1.22 (0.80 to 1.85) |  | 0.205 | 1.11 (0.94 to 1.32) |
| **Distal** |  |  |  |  |  |  |  |
| *N*<sub>cases</sub> | 230 | 229 | 59 | 33 |  |  |  |
| Multivariable | 1 | 0.94 (0.77 to 1.15) | 1.36 (1.00 to 1.86) | 1.21 (0.79 to 1.88) |  | 0.157 | 1.13 (0.95 to 1.33) |
| **Rectal** |  |  |  |  |  |  |  |
| *N*<sub>cases</sub> | 97 | 77 | 29 | 20 |  |  |  |
| Multivariable | 1 | 0.79 (0.57 to 1.09) | 1.80 (1.14 to 2.84) | 2.19 (1.20 to 4.00) |  | 0.001 | 1.48 (1.16 to 1.88) |
|  | Total fructose intake during adolescence, % of calorie |  |  |  |  |  | Per 5% of<br>calorie/day<br>increase |
|  | Q1<br>(<7.9) | Q2<br>(7.9–<9.2) | Q3<br>(9.2–<10.3) | Q4<br>(10.3–<11.8) | Q5<br>(≥11.8) | P value<br>for trend |  |
| **Low-risk adenoma** |  |  |  |  |  |  |  |
| *N*<sub>cases</sub>/*N*<sub>controls</sub> | 301/9795 | 267/9807 | 296/9765 | 249/9776 | 271/9809 |  |  |
| Multivariable | 1 | 0.92 (0.77 to 1.09) | 1.05 (0.87 to 1.26) | 0.91 (0.75 to 1.11) | 1.02 (0.83 to 1.26) | 0.821 | 1.02 (0.86 to 1.20) |
| **High-risk adenoma** |  |  |  |  |  |  |  |
| *N*<sub>cases</sub> | 191 | 213 | 214 | 242 | 222 |  |  |
| Multivariable | 1 | 1.20 (0.98 to 1.48) | 1.27 (1.02 to 1.57) | 1.53 (1.23 to 1.92) | 1.44 (1.14 to 1.83) | 0.001 | 1.36 (1.14 to 1.62) |
| **Proximal** |  |  |  |  |  |  |  |
| *N*<sub>cases</sub> | 129 | 120 | 122 | 138 | 118 |  |  |
| Multivariable | 1 | 1.02 (0.79 to 1.33) | 1.11 (0.84 to 1.46) | 1.37 (1.03 to 1.82) | 1.23 (0.91 to 1.67) | 0.064 | 1.25 (0.99 to 1.58) |
| **Distal** |  |  |  |  |  |  |  |
| *N*<sub>cases</sub> | 94 | 99 | 111 | 121 | 126 |  |  |
| Multivariable | 1 | 1.12 (0.84 to 1.50) | 1.31 (0.97 to 1.77) | 1.50 (1.11 to 2.03) | 1.54 (1.11 to 2.13) | 0.004 | 1.44 (1.12 to 1.84) |
| **Rectal** |  |  |  |  |  |  |  |
| *N*<sub>cases</sub> | 33 | 49 | 34 | 58 | 49 |  |  |
| Multivariable | 1 | 1.74 (1.09 to 2.77) | 1.28 (0.77 to 2.13) | 2.45 (1.49 to 4.02) | 2.08 (1.23 to 3.53) | 0.004 | 1.74 (1.19 to 2.54) |

\*Low-risk: small (<1cm), tubular, and single adenoma, high-risk: large ( $\geq 1$ cm), any villous histology, high-grade dysplasia, or more than 2 adenomas.
†N, number of endoscopies; due to multiple endoscopies during follow-up per each participant, N is larger than number of participants.
‡Adjusted for age, time period of endoscopy, number of endoscopies during the study period (continuous), time since most recent endoscopy (continuous), and reason for current endoscopy, family history of colorectal cancer, menopausal status/menopausal hormone use, current aspirin use $\geq 2$ times/wk, history of type 2 diabetes, adult height (continuous), BMI at age 18 y, current BMI, smoking status at 19 y, current smoking status, alcohol intake at 18–22 y, current alcohol intake, physical activity during grades 9–12, current physical activity, adolescent and current dietary intake (total calorie, total calcium, vitamin D, total folate, total fiber, fruits, vegetables, and dairy), current total red meat intake, a western dietary pattern score during adolescence, and current sugar-sweetened beverage or total fructose intake.

### Sensitivity analysis

Overall, sensitivity analysis results were consistent with the principal findings. In brief, further adjustment for ASB and fruit juice intake or prudent dietary pattern did not materially change the associations of SSB and fructose intake with adenoma risk (supplementary table 5). When energy-adjusted intake of sugars was examined instead of nutrient densities, results were similar (supplementary table 6). An alternative definition of high-risk adenoma (≥1 cm, any villous histology, high-grade dysplasia, or ≥3 adenomas)^48^ yielded similar results, especially for high-risk rectal adenoma (supplementary table 7). Results for proximal adenoma were similar when restricting analyses to women who underwent colonoscopy (after excluding those with sigmoidoscopy only; supplementary table 8). After additional adjustment for glycemic index and glycemic load, potential mediators, positive associations were substantially attenuated, especially after adjustment for glycemic load: per 5% increase in calorie/day of total fructose, odd ratios were 1.08 for total and 1.24 for high-risk adenoma (supplementary table 5).

### Stratified analysis

Associations of SSB and fructose intake with adenoma risk did not differ appreciably by family history of CRC, adolescent BMI, physical activity, smoking, or alcohol consumption (all P≥0.15 for interaction; fig 1, supplementary tables 9 and 10). When stratified by age at diagnosis (<55 *v* ≥55 years), positive associations were similar across strata although the majority of cases were diagnosed at relatively young ages (76.5% before 55 years, 99.6% before 65 years) (supplementary table 11).

**Fig 1.**
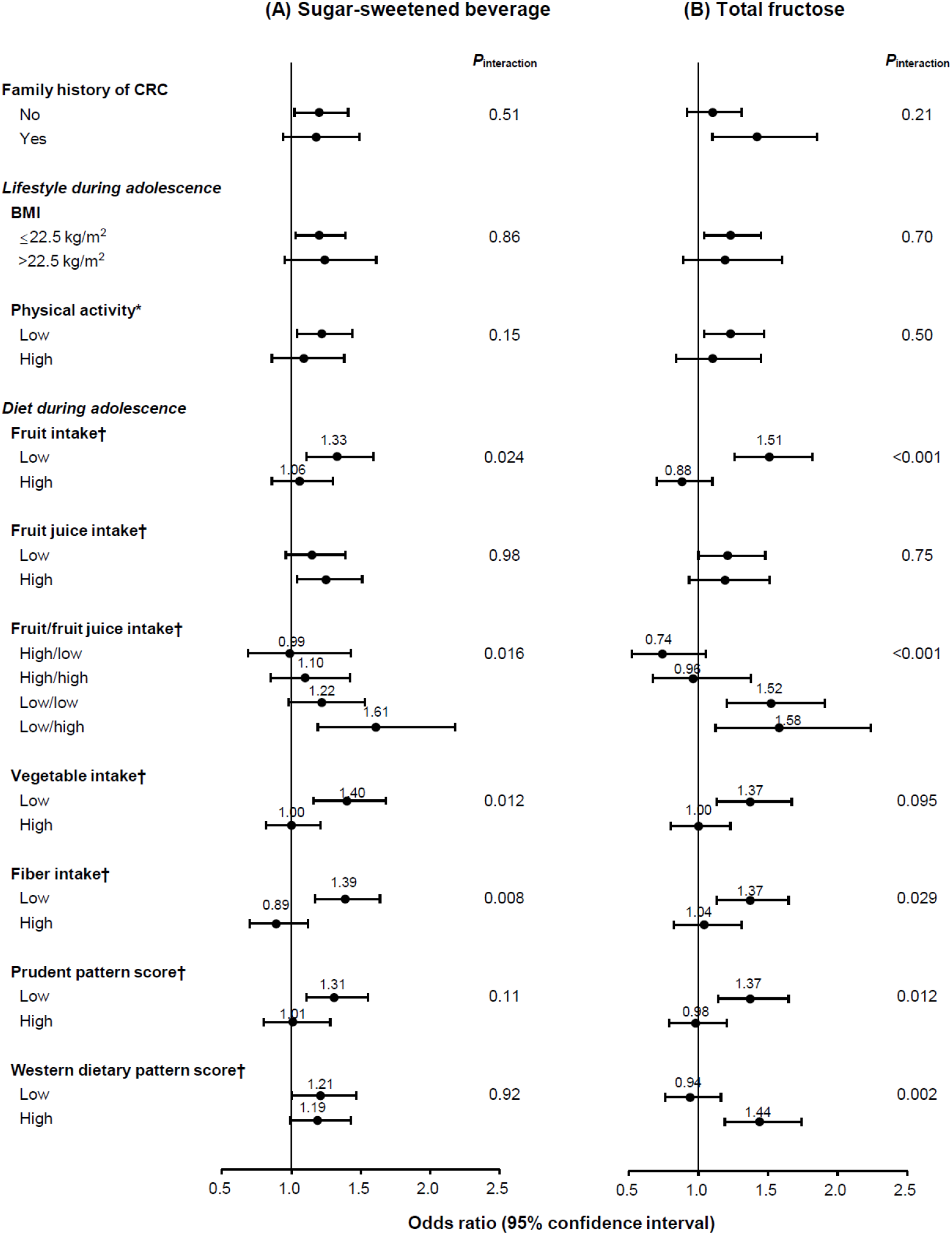
**Multivariable odds ratios and 95% confidence intervals of total colorectal adenoma according to (A) sugar-sweetened beverage and (B) total fructose intake during adolescence by family history, lifestyle, and dietary factors in the Nurses’ Health Study II, 1998-2015** CRC, colorectal cancer; BMI, body mass index Data were adjusted for age, time period of endoscopy, number of endoscopies during the study period (continuous), time since most recent endoscopy (continuous), and reason for current endoscopy, family history of colorectal cancer, menopausal status/menopausal hormone use, current aspirin use ≥2 times/wk, history of type 2 diabetes, adult height (continuous), BMI at age 18 y, current BMI, smoking status at 19 y, current smoking status, alcohol intake at 18–22 y, current alcohol intake, physical activity during grades 9–12, current physical activity, adolescent and current dietary intake (total calorie, total calcium, vitamin D, total folate, total fiber, fruits, vegetables, and dairy), current total red meat intake, a western dietary pattern score during adolescence, and current sugar-sweetened beverage or total fructose intake except for the stratifying variable of each stratum. (A) Comparison of sugar-sweetened beverage intake between categories of ≥1 serving/d vs. <1 serving/wk (referent). (B) Comparison of total fructose intake between the highest vs. lowest (referent) quintile. *High physical activity was defined as the highest tertile (≥59 MET-hr/wk); low physical activity as the 2 lower tertiles (<59 MET-hr/wk). †Cut-off values were median intake values (fruits, 1.3 serving/d; fruit juice, 0.4 serving/d; vegetables, 2.8 serving/d).

Positive associations between sugar intake and adenoma risk were significantly stronger among women with low fruit intake (<1.3 servings/day) during adolescence compared to those with high intake (≥1.3 servings/day). Among women with low fruit intake, odds ratios of total adenoma were 1.33 (95% confidence interval 1.11 to 1.59; P=0.024 for interaction) for SSBs (≥1 serving/day *v* <1 serving/week) and 1.51 (1.26 to 1.82; P<0.001 for interaction) for total fructose (highest *v* lowest quintile). Similar differential associations were observed after stratification by vegetable and fiber intake, and prudent dietary pattern. In contrast, positive associations with adenoma risk did not differ appreciably by fruit juice intake (P≥0.75 for interaction). Associations were further examined by joint categories of fruit (high/low) and fruit juice (high/low) intake. Positive associations were stronger in the ‘low fruit/high fruit juice’ subgroup (odds ratio, 1.61 for SSBs, 1.58 for total fructose) with significant differences across subgroups (P≤0.016 for interaction; fig 1). Stratified analysis results for high-risk adenoma were almost identical to those for total adenoma (supplementary table 10).

### Joint analysis of adolescent and adult diet

Compared to women with low SSB or fructose intake during both adolescence and adulthood, women with high intake during adolescence had increased risk of total, rectal, and high-risk adenoma (supplementary fig 1, supplementary table 12). Associations did not differ significantly between the ‘high adolescent/high adult intake’ and ‘high adolescent/low adult intake’ groups. However, these results need to be interpreted with caution because of higher added sugar and calorie intake during adolescence and differences in metabolism and nutritional/caloric requirements between adolescents and adults.

### Substitution analysis

The 2015–2020 Dietary Guidelines for Americans recommend 2 cup-equivalents of fruits (whole fruits and 100% fruit juice) at the 2000-calorie level and 2–3 cup-equivalents of dairy per day for children and adolescents.^49^ Substituting 1 serving/day of fruit juice for 1 serving/day of SSBs during adolescence was not associated with lower risk of adenoma (supplementary table 13). In contrast, replacement with 2 servings/day of fruits for 2 servings/day of SSBs was marginally associated with reduced risk of proximal (odds ratio 0.75, 95% confidence interval 0.54 to 1.05) and rectal (0.62, 0.35 to 1.08) adenoma. Substituting 2 servings/day of dairy products for 2 servings/day of SSBs was associated with lower risk of total (0.82, 0.64 to 1.05) and rectal (0.53, 0.29 to 0.94) adenoma.

## DISCUSSION

### Principal findings

In this large cohort of young women, independent of adult intake, higher intake of SSBs and sugars during adolescence was significantly associated with increased risk of total and high-risk adenoma, particularly high-risk rectal adenoma. Results were similar, albeit slightly weaker, for added sugar and total sugar. Neither SSB nor sugar intake was associated with risk of serrated lesions. Thus, high sugar intake during adolescence may be etiologically more important for CRC arising from the conventional adenoma-carcinoma sequence, which accounts for approximately 85% of CRC,^2^ rather than those originating from the serrated neoplasia pathway.

### Comparison with other studies

Prospective studies on adult SSB or sugar intake in relation to CRC risk have generally found null associations, including 2 comprehensive pooled analyses of prospective studies as well as a recent large cohort study.^18 23 24^ We also did not observe significant associations of adult SSB and sugar intake with adenoma risk. In 2018, the World Cancer Research Fund/American Institute for Cancer Research reported that evidence for sugars and foods containing sugars with regard to CRC risk was limited.^4 24^ However, this conclusion was based on intake during adulthood, mostly capturing mid-to-late adulthood cases. In contrast, little is known about the influence of early-life diet on colorectal carcinogenesis. Emerging evidence has suggested a potential role for adolescent diet in CRC development. For example, severe energy restriction during adolescence was associated with lower risk of CRC and persistent epigenetic changes involved in colorectal carcinogenesis.^50 51^ In the NHSII, we previously reported that a western dietary pattern and physical inactivity during adolescence were associated with higher risk of adenoma, whereas red and processed meat intake, established CRC risk factors in adulthood, was not associated.^52-54^

To our knowledge, no previous study has investigated the association between adolescent sugar intake and colorectal neoplasia. During adolescence, accompanied by accelerated cell proliferation, distinctive hormonal and metabolic changes occur, including physiological (obesity-unrelated) hyperinsulinemia, decreased insulin sensitivity, and elevated IGF1 levels (up to 4-fold higher than in adulthood).^27^ Therefore, adolescence might be a critical developmental period of enhanced susceptibility to high sugar intake, further adversely affecting insulin sensitivity. Our joint analysis results indicated that higher risk of adenoma was influenced by sugar intake primarily during adolescence, but not during adulthood.

### Meaning of the study

Several biological mechanisms may explain our findings. First, hyperinsulinemia and insulin resistance can play important roles. The high amount of liquid sugar in SSBs can induce rapid spikes in blood glucose and insulin levels, which over time lead to insulin resistance and elevated free IGF1 levels.^10^ The insulin/IGF1 system can promote carcinogenesis by activating intracellular signaling pathways related to altered gene expression, stimulating cell proliferation, differentiation, and angiogenesis, and inhibiting apoptosis.^3 55 56^ We found that additional adjustment for dietary glycemic load substantially attenuated positive associations of high sugar intake, supporting this hypothesis.

Second, hyperglycemia may exacerbate chronic inflammation that has been implicated in CRC pathogenesis.^57^ Previous studies have reported that SSB intake was significantly associated with increased levels of circulating inflammatory cytokines and biomarkers (e.g., C-reactive protein, interleukin-6, tumor necrosis factor receptors).^17 58^ Third, the distinctive metabolism of fructose, a major ingredient of SSBs, can exert additional adverse effects. Unlike glucose, fructose is metabolized predominantly in the liver after absorption in the small intestine.^10^ When fructose intake chronically exceeds the metabolic capacity of the liver, fructose triggers hepatic *de novo* lipogenesis, promoting visceral and ectopic fat accumulation, glucose intolerance, and insulin resistance.^9 59 60^ Additionally, a recent experimental animal study found that fructose was metabolized into glucose in murine small intestine as well; and intestinal fructose-to-glucose conversion was not suppressed by insulin, implying a novel unregulated pathway.^61^

Finally, fructose may affect carcinogenesis by directly acting on colorectal cells or interacting with the gut microbiome. Although fructose is readily absorbed in the small intestine, high doses or rapid flux of fructose could saturate small intestine clearance capacity, with excess fructose reaching the colon.^61 62^ An 8-week oral administration of high-fructose corn syrup in mice enhanced colorectal tumor cell growth, even at a moderate dose, in the absence of obesity and metabolic syndrome, implying direct effects of fructose on tumor cell metabolism.^63^ In addition, sugars may change the gut microbiome composition,^64 65^ which could affect CRC development via modulation of gut immune and metabolic responses, and epigenetic alterations.^66 67^

In stratified analyses, positive associations of high sugar intake were significantly stronger among women with low fruit, vegetable, or fiber intake during adolescence than those with high intake. Unlike fruits or vegetables, fruit juice did not appear to offset the adverse effects of high sugar intake. Moreover, positive associations of high sugar intake were stronger among women consuming low fruits/high fruit juice during adolescence compared to those consuming high fruits/low fruit juice. We found no benefits of substituting fruit juice for SSBs. These results may be explained as follows: although fruits and some vegetables contain naturally occurring sugars,^34^ many beneficial micronutrients and potential anti-tumorigenic agents (e.g., fiber, folate, vitamins) may offset or dilute the adverse effects of sugars.^4^ Furthermore, one of the key differences between whole fruits and fruit juice may be intestinal fructose release rates.^61^ Fructose in whole fruits is slowly digested due to fiber content and the need to disrupt cell structure, facilitating gradual and complete intestinal clearance.^10^ In contrast, rapid flux of liquid fructose from fruit juice may exceed small intestine uptake capacity, resulting in fructose overflow to the liver and colon.^10 61 62^

We also found stronger associations among women with unhealthy (low prudent and high western) dietary patterns during adolescence than those with healthy patterns. These results suggest that potential additive and interactive effects of foods consumed together in an overall diet may influence the biological responses of high sugar intake.^68^ While excessive sugar intake may elicit unfavorable postprandial glucose and insulin responses, cumulative effects of acute hyperinsulinemia can contribute to colorectal carcinogenesis, particularly when combined with overall unhealthy dietary patterns that affect underlying chronic insulin resistance.^68 69^

### Strengths and limitations of the study

This study has several strengths. To our knowledge, this is the first prospective study investigating the role of high sugar intake during adolescence in colorectal neoplasia. The large sample size of 33106 women and 4744 CRC precursor cases enabled investigating various subtypes and subsites and conducting stratified analyses. Diet and lifestyle information was validated and obtained throughout different life stages, enabling us to examine both independent effects and joint associations of adolescent and adult diet. We comprehensively controlled for various known and purported CRC risk factors during both adolescence and adulthood. A majority of CRC precursors were diagnosed at relatively young ages (77% were under 55 years), supporting a plausible link between early-life exposure and early steps of colorectal carcinogenesis. Moreover, in rigorous sensitivity analyses, the principal results were robust.

Potential limitations of this study need to be considered. First, measurement error in dietary assessment can never be excluded when using self-reported questionnaire data. However, the HS-FFQ showed reasonable reproducibility and validity.^29 31^ Also, non-differential misclassification of exposure generally attenuates associations towards the null. Second, residual and unmeasured confounding could exist. High sugar intake could be a marker for a generally unhealthy diet or lifestyle that might track throughout life. However, we controlled for numerous dietary and lifestyle factors as well as overall dietary patterns during both adolescence and adulthood. Finally, the study population consisted of predominantly white female nurses and results may not be generalizable to other populations. However, exposure-CRC associations in our cohorts have been highly consistent with findings in diverse populations,^4 53 70-72^ suggesting a common underlying biology.

### Policy implications

If confirmed, our findings may have substantial public health implications for the primary prevention of CRC. In many high-income countries, the rising incidence of early-onset CRC has been primarily driven by a disproportional increase in distal and rectal cancers,^3 5^ known to arise mainly through the adenoma-carcinoma pathway.^73^ The majority of early-onset CRC is sporadic and the causes remain unclear.^3 73^ However, the observed birth cohort effect strongly suggests that early-life exposures are involved and causes likely stem from changes in diet and lifestyle factors in youths.^5^ In our results, associations of high sugar intake were stronger for distally-located high-risk adenoma, especially high-risk rectal adenoma; and substituting dairy products for SSBs was associated with lower risk of rectal adenoma. In the early 1970s, the national mean intake of SSBs among U.S. adolescents was higher than in our participants,^47^ and recent data on global adolescent SSB consumption have indicated a marked increase.^10 11 13^ Therefore, if applied to the current general population, the effect size may be even larger than observed in our results. Our findings suggest that adolescence can be a critical developmental period of enhanced susceptibility to high sugar intake, supporting the importance of limiting sugar intake and replacing SSBs with healthy alternatives during early-life in an effort to reduce risk of CRC precursors and possibly CRC.

## Conclusions

In conclusion, SSB and sugar intake during adolescence was significantly associated with increased risk of total and high-risk adenoma, especially high-risk rectal adenoma, but not serrated lesions. Thus, high sugar intake during early-life may promote precancerous lesions in CRC, primarily through the adenoma-carcinoma pathway. Future studies are needed to determine whether high sugar intake during adolescence contributes to CRC risk, particularly the recent upward trend in early-onset CRC.

## Data Availability

Data Sharing: No additional data available.

## Acknowledgments

The authors would like to thank the Channing Division of Network Medicine, Department of Medicine, Brigham and Women’s Hospital and Harvard Medical School. The authors would like to thank the participants and staff of the Nurses’ Health Study II for their valuable contributions as well as the following state cancer registries for their help: AL, AZ, AR, CA, CO, CT, DE, FL, GA, ID, IL, IN, IA, KY, LA, ME, MD, MA, MI, NE, NH, NJ, NY, NC, ND, OH, OK, OR, PA, RI, SC, TN, TX, VA, WA and WY. The authors assume full responsibility for analyses and interpretation of these data.

## Ethical approval

The study protocol was approved by the institutional review boards of the Brigham and Women’s Hospital and Harvard T.H. Chan School of Public Health, and those of participating registries as required. Return of the mailed questionnaire was considered to imply informed consent.

## Transparency statement

Data described in the manuscript, code book, and analytic code will not be made publicly available. Further information including the procedures for obtaining and accessing data from the Nurses’ Health Studies II is described at https://www.nurseshealthstudy.org/researchers.

The lead author (H-KJ) affirms that the manuscript is an honest, accurate, and transparent account of the study being reported; that no important aspects of the study have been omitted; and that any discrepancies from the study as originally planned have been explained.

## Dissemination declaration

We have reported whether we plan to disseminate the results to study participants and or patient organisations.

## Data Sharing

No additional data available.

## Licence Statement

This is an open access article distributed in accordance with the Creative Commons Attribution Non Commercial (CC BY-NC 4.0) license, which permits others to distribute, remix, adapt, build upon this work non-commercially, and license their derivative works on different terms, provided the original work is properly cited and the use is non-commercial. See: http://creativecommons.org/licenses/by-nc/4.0.

The Corresponding Author has the right to grant on behalf of all authors and does grant on behalf of all authors, an exclusive licence on a worldwide basis to the BMJ Publishing Group Ltd to permit this article (if accepted) to be published in BMJ editions and any other BMJPGL products and sublicences such use and exploit all subsidiary rights, as set out in our licence.

## References

1. Arnold M, Sierra MS, Laversanne M, et al. Global patterns and trends in colorectal cancer incidence and mortality. Gut 2017;66(4):683–91. doi: 10.1136/gutjnl-2015-310912 [published Online First: 2016/01/29]

2. Strum WB. Colorectal Adenomas. N Engl J Med 2016;374(11):1065–75. doi: 10.1056/NEJMra1513581 [published Online First: 2016/03/18]

3. Keum N, Giovannucci E. Global burden of colorectal cancer: emerging trends, risk factors and prevention strategies. Nat Rev Gastroenterol Hepatol 2019;16(12):713–32. doi: 10.1038/s41575-019-0189-8 [published Online First: 2019/08/29]

4. Research. WCRFAIfC. Diet, Nutrition, Physical Activity and Cancer: a Global Perspective. Continuous Update Project Expert Report 2018. Available at dietandcancerreport.org.

5. Siegel RL, Miller KD, Goding Sauer A, et al. Colorectal cancer statistics, 2020. CA Cancer J Clin 2020;70(3):145–64. doi: 10.3322/caac.21601 [published Online First: 2020/03/07]

6. Siegel RL, Jakubowski CD, Fedewa SA, et al. Colorectal Cancer in the Young: Epidemiology, Prevention, Management. Am Soc Clin Oncol Educ Book 2020;40:1–14. doi: 10.1200/edbk_279901 [published Online First: 2020/04/22]

7. Yang Q, Zhang Z, Gregg EW, et al. Added sugar intake and cardiovascular diseases mortality among US adults. JAMA Intern Med 2014;174(4):516–24. doi: 10.1001/jamainternmed.2013.13563 [published Online First: 2014/02/05]

8. Malik VS, Li Y, Pan A, et al. Long-Term Consumption of Sugar-Sweetened and Artificially Sweetened Beverages and Risk of Mortality in US Adults. Circulation 2019;139(18):2113–25. doi: 10.1161/circulationaha.118.037401 [published Online First: 2019/03/19]

9. Bray GA, Nielsen SJ, Popkin BM. Consumption of high-fructose corn syrup in beverages may play a role in the epidemic of obesity. Am J Clin Nutr 2004;79(4):537–43. doi: 10.1093/ajcn/79.4.537 [published Online First: 2004/03/31]

10. Malik VS, Hu FB. Sugar-Sweetened Beverages and Cardiometabolic Health: An Update of the Evidence. Nutrients 2019;11(8) doi: 10.3390/nu11081840 [published Online First: 2019/08/11]

11. Yang L, Bovet P, Liu Y, et al. Consumption of Carbonated Soft Drinks Among Young Adolescents Aged 12 to 15 Years in 53 Low- and Middle-Income Countries. Am J Public Health 2017;107(7):1095–100. doi: 10.2105/ajph.2017.303762 [published Online First: 2017/05/19]

12. Rosinger A, Herrick K, Gahche J, et al. Sugar-sweetened Beverage Consumption Among U.S. Adults, 2011-2014. NCHS Data Brief 2017(270):1–8. [published Online First: 2017/01/31]

13. Rosinger A, Herrick K, Gahche J, et al. Sugar-sweetened Beverage Consumption Among U.S. Youth, 2011-2014. NCHS Data Brief 2017(271):1–8. [published Online First: 2017/01/31]

14. Makarem N, Bandera EV, Nicholson JM, et al. Consumption of Sugars, Sugary Foods, and Sugary Beverages in Relation to Cancer Risk: A Systematic Review of Longitudinal Studies. Annu Rev Nutr 2018;38:17–39. doi: 10.1146/annurev-nutr-082117-051805 [published Online First: 2018/05/29]

15. Michaud DS, Fuchs CS, Liu S, et al. Dietary glycemic load, carbohydrate, sugar, and colorectal cancer risk in men and women. Cancer Epidemiol Biomarkers Prev 2005;14(1):138–47. [published Online First: 2005/01/26]

16. Coleman HG, Ness RM, Smalley WE, et al. Aspects of dietary carbohydrate intake are not related to risk of colorectal polyps in the Tennessee Colorectal Polyp Study. Cancer Causes Control 2015;26(8):1197–202. doi: 10.1007/s10552-015-0605-5 [published Online First: 2015/06/10]

17. Yu Z, Ley SH, Sun Q, et al. Cross-sectional association between sugar-sweetened beverage intake and cardiometabolic biomarkers in US women. Br J Nutr 2018;119(5):570–80. doi: 10.1017/s0007114517003841 [published Online First: 2018/03/07]

18. Pacheco LS, Anderson CAM, Lacey JV, Jr., et al. Sugar-sweetened beverages and colorectal cancer risk in the California Teachers Study. PLoS One 2019;14(10):e0223638. doi: 10.1371/journal.pone.0223638 [published Online First: 2019/10/10]

19. Giovannucci E. Insulin, insulin-like growth factors and colon cancer: a review of the evidence. J Nutr 2001;131(11 Suppl):3109s–20s. doi: 10.1093/jn/131.11.3109S [published Online First: 2001/11/06]

20. Schoen RE, Tangen CM, Kuller LH, et al. Increased blood glucose and insulin, body size, and incident colorectal cancer. J Natl Cancer Inst 1999;91(13):1147–54. doi: 10.1093/jnci/91.13.1147 [published Online First: 1999/07/07]

21. Kaaks R, Toniolo P, Akhmedkhanov A, et al. Serum C-peptide, insulin-like growth factor (IGF)-I, IGF-binding proteins, and colorectal cancer risk in women. J Natl Cancer Inst 2000;92(19):1592–600. doi: 10.1093/jnci/92.19.1592 [published Online First: 2000/10/06]

22. Ma J, Giovannucci E, Pollak M, et al. A prospective study of plasma C-peptide and colorectal cancer risk in men. J Natl Cancer Inst 2004;96(7):546–53. doi: 10.1093/jnci/djh082 [published Online First: 2004/04/08]

23. Zhang X, Albanes D, Beeson WL, et al. Risk of colon cancer and coffee, tea, and sugar-sweetened soft drink intake: pooled analysis of prospective cohort studies. J Natl Cancer Inst 2010;102(11):771–83. doi: 10.1093/jnci/djq107 [published Online First: 2010/05/11]

24. Project. ICLCU. World Cancer Research Fund International Systematic Literature Review: The Associations between Food, Nutrition and Physical Activity and the Risk of Colorectal Cancer, 2017.

25. Potischman N, Linet MS. Invited commentary: are dietary intakes and other exposures in childhood and adolescence important for adult cancers? Am J Epidemiol 2013;178(2):184–9. doi: 10.1093/aje/kwt101 [published Online First: 2013/06/25]

26. Nimptsch K, Wu K. Is Timing Important? The Role of Diet and Lifestyle during Early Life on Colorectal Neoplasia. Curr Colorectal Cancer Rep 2018;14(1):1–11. doi: 10.1007/s11888-018-0396-7 [published Online First: 2018/08/25]

27. Hannon TS, Janosky J, Arslanian SA. Longitudinal study of physiologic insulin resistance and metabolic changes of puberty. Pediatr Res 2006;60(6):759–63. doi: 10.1203/01.pdr.0000246097.73031.27 [published Online First: 2006/10/27]

28. Bao Y, Bertoia ML, Lenart EB, et al. Origin, Methods, and Evolution of the Three Nurses’ Health Studies. Am J Public Health 2016;106(9):1573–81. doi: 10.2105/ajph.2016.303338 [published Online First: 2016/07/28]

29. Maruti SS, Feskanich D, Colditz GA, et al. Adult recall of adolescent diet: reproducibility and comparison with maternal reporting. American journal of epidemiology 2005;161(1):89–97. doi: 10.1093/aje/kwi019

30. Nurses’ Health Study II. Special High School Questionnaire 1998 [Available from: https://www.nurseshealthstudy.org/sites/default/files/questionnaires/1998.nhs2.hs_diet.pdf.

31. Maruti SS, Feskanich D, Rockett HR, et al. Validation of adolescent diet recalled by adults. Epidemiology (Cambridge, Mass) 2006;17(2):226–29. doi: 10.1097/01.ede.0000198181.86685.49

32. Hu FB, Rimm E, Smith-Warner SA, et al. Reproducibility and validity of dietary patterns assessed with a food-frequency questionnaire. Am J Clin Nutr 1999;69(2):243–9. doi: 10.1093/ajcn/69.2.243 [published Online First: 1999/02/16]

33. Feskanich D, Rimm EB, Giovannucci EL, et al. Reproducibility and validity of food intake measurements from a semiquantitative food frequency questionnaire. J Am Diet Assoc 1993;93(7):790–6. doi: 10.1016/0002-8223(93)91754-e [published Online First: 1993/07/01]

34. Johnson RK, Appel LJ, Brands M, et al. Dietary sugars intake and cardiovascular health: a scientific statement from the American Heart Association. Circulation 2009;120(11):1011–20. doi: 10.1161/circulationaha.109.192627 [published Online First: 2009/08/26]

35. US Department of Agriculture. Composition of foods: raw, processed, and prepared, 1963–1980. (Agricultural handbook no. 8). Washington, DC: US Department of Agriculture, 1980.

36. Willett W, Stampfer MJ. Total energy intake: implications for epidemiologic analyses. Am J Epidemiol 1986;124(1):17–27. doi: 10.1093/oxfordjournals.aje.a114366 [published Online First: 1986/07/01]

37. Willett WC. Nutritional Epidemiology, Second Edition. New York: Oxford University Press 1998.

38. Willett W. Nutritional Epidemiology. New York: Oxford University Press USA 2013.

39. Hu FB. Dietary pattern analysis: a new direction in nutritional epidemiology. Curr Opin Lipidol 2002;13(1):3–9. doi: 10.1097/00041433-200202000-00002 [published Online First: 2002/01/16]

40. American Cancer Society. About Colorectal Cancer 2018 [Available from: https://www.cancer.org/cancer/colon-rectal-cancer/about/what-is-colorectal-cancer.html May 03, 2020.

41. East JE, Vieth M, Rex DK. Serrated lesions in colorectal cancer screening: detection, resection, pathology and surveillance. Gut 2015;64(6):991–1000. doi: 10.1136/gutjnl-2014-309041 [published Online First: 2015/03/10]

42. Andersen PK, Gill RD. Cox’s Regression Model for Counting Processes: A Large Sample Study.

43. He X, Wu K, Ogino S, et al. Association Between Risk Factors for Colorectal Cancer and Risk of Serrated Polyps and Conventional Adenomas. Gastroenterology 2018;155(2):355-73.e18. doi: 10.1053/j.gastro.2018.04.019 [published Online First: 2018/04/28]

44. Bernstein AM, de Koning L, Flint AJ, et al. Soda consumption and the risk of stroke in men and women. Am J Clin Nutr 2012;95(5):1190–9. doi: 10.3945/ajcn.111.030205 [published Online First: 2012/04/12]

45. Bernstein AM, Sun Q, Hu FB, et al. Major dietary protein sources and risk of coronary heart disease in women. Circulation 2010;122(9):876–83. doi: 10.1161/circulationaha.109.915165 [published Online First: 2010/08/18]

46. Halton TL, Willett WC, Liu S, et al. Potato and french fry consumption and risk of type 2 diabetes in women. The American journal of clinical nutrition 2006;83(2):284–90. [published Online First: 2006/02/14]

47. Chun OK, Chung CE, Wang Y, et al. Changes in intakes of total and added sugar and their contribution to energy intake in the U.S. Nutrients 2010;2(8):834–54. doi: 10.3390/nu2080834 [published Online First: 2010/08/01]

48. Gupta S, Lieberman D, Anderson JC, et al. Recommendations for Follow-Up After Colonoscopy and Polypectomy: A Consensus Update by the US Multi-Society Task Force on Colorectal Cancer. Gastroenterology 2020;158(4):1131-53.e5. doi: 10.1053/j.gastro.2019.10.026 [published Online First: 2020/02/12]

49. U.S. Department of Health and Human Services and U.S. Department of Agriculture. 2015–2020 Dietary Guidelines for Americans. 8th Edition. December 2015. Available at http://health.gov/dietaryguidelines/2015/guidelines/.

50. Hughes LA, van den Brandt PA, de Bruïne AP, et al. Early life exposure to famine and colorectal cancer risk: a role for epigenetic mechanisms. PLoS One 2009;4(11):e7951. doi: 10.1371/journal.pone.0007951 [published Online First: 2009/12/04]

51. Hughes LA, van den Brandt PA, Goldbohm RA, et al. Childhood and adolescent energy restriction and subsequent colorectal cancer risk: results from the Netherlands Cohort Study. Int J Epidemiol 2010;39(5):1333–44. doi: 10.1093/ije/dyq062 [published Online First: 2010/04/30]

52. Nimptsch K, Malik VS, Fung TT, et al. Dietary patterns during high school and risk of colorectal adenoma in a cohort of middle-aged women. Int J Cancer 2014;134(10):2458–67. doi: 10.1002/ijc.28578 [published Online First: 2014/02/05]

53. Rezende LFM, Lee DH, Keum N, et al. Physical activity during adolescence and risk of colorectal adenoma later in life: results from the Nurses’ Health Study II. Br J Cancer 2019;121(1):86–94. doi: 10.1038/s41416-019-0454-1 [published Online First: 2019/05/23]

54. Nimptsch K, Bernstein AM, Giovannucci E, et al. Dietary intakes of red meat, poultry, and fish during high school and risk of colorectal adenomas in women. Am J Epidemiol 2013;178(2):172–83. doi: 10.1093/aje/kwt099 [published Online First: 2013/06/21]

55. Tran TT, Medline A, Bruce WR. Insulin promotion of colon tumors in rats. Cancer Epidemiol Biomarkers Prev 1996;5(12):1013–5. [published Online First: 1996/12/01]

56. Vigneri PG, Tirrò E, Pennisi MS, et al. The Insulin/IGF System in Colorectal Cancer Development and Resistance to Therapy. Front Oncol 2015;5:230. doi: 10.3389/fonc.2015.00230 [published Online First: 2015/11/04]

57. Wu J, Cai Q, Li H, et al. Circulating C-reactive protein and colorectal cancer risk: a report from the Shanghai Men’s Health Study. Carcinogenesis 2013;34(12):2799–803. doi: 10.1093/carcin/bgt288 [published Online First: 2013/08/30]

58. de Koning L, Malik VS, Kellogg MD, et al. Sweetened beverage consumption, incident coronary heart disease, and biomarkers of risk in men. Circulation 2012;125(14):1735-41, s1. doi: 10.1161/circulationaha.111.067017 [published Online First: 2012/03/14]

59. Malik VS, Hu FB. Fructose and Cardiometabolic Health: What the Evidence From Sugar-Sweetened Beverages Tells Us. J Am Coll Cardiol 2015;66(14):1615–24. doi: 10.1016/j.jacc.2015.08.025 [published Online First: 2015/10/03]

60. Wu T, Giovannucci E, Pischon T, et al. Fructose, glycemic load, and quantity and quality of carbohydrate in relation to plasma C-peptide concentrations in US women. Am J Clin Nutr 2004;80(4):1043–9. doi: 10.1093/ajcn/80.4.1043 [published Online First: 2004/09/28]

61. Jang C, Hui S, Lu W, et al. The Small Intestine Converts Dietary Fructose into Glucose and Organic Acids. Cell Metab 2018;27(2):351-61.e3. doi: 10.1016/j.cmet.2017.12.016 [published Online First: 2018/02/08]

62. Zhao S, Jang C, Liu J, et al. Dietary fructose feeds hepatic lipogenesis via microbiota-derived acetate. Nature 2020;579(7800):586–91. doi: 10.1038/s41586-020-2101-7 [published Online First: 2020/03/28]

63. Goncalves MD, Lu C, Tutnauer J, et al. High-fructose corn syrup enhances intestinal tumor growth in mice. Science 2019;363(6433):1345–49. doi: 10.1126/science.aat8515 [published Online First: 2019/03/23]

64. Do MH, Lee E, Oh MJ, et al. High-Glucose or -Fructose Diet Cause Changes of the Gut Microbiota and Metabolic Disorders in Mice without Body Weight Change. Nutrients 2018;10(6) doi: 10.3390/nu10060761 [published Online First: 2018/06/15]

65. Di Rienzi SC, Britton RA. Adaptation of the Gut Microbiota to Modern Dietary Sugars and Sweeteners. Adv Nutr 2020;11(3):616–29. doi: 10.1093/advances/nmz118 [published Online First: 2019/11/07]

66. Wong SH, Yu J. Gut microbiota in colorectal cancer: mechanisms of action and clinical applications. Nat Rev Gastroenterol Hepatol 2019;16(11):690–704. doi: 10.1038/s41575-019-0209-8 [published Online First: 2019/09/27]

67. Song M, Chan AT, Sun J. Influence of the Gut Microbiome, Diet, and Environment on Risk of Colorectal Cancer. Gastroenterology 2020;158(2):322–40. doi: 10.1053/j.gastro.2019.06.048 [published Online First: 2019/10/07]

68. Giovannucci E. A framework to understand diet, physical activity, body weight, and cancer risk. Cancer Causes Control 2018;29(1):1–6. doi: 10.1007/s10552-017-0975-y [published Online First: 2017/11/11]

69. Tabung FK, Wang W, Fung TT, et al. Association of dietary insulinemic potential and colorectal cancer risk in men and women. Am J Clin Nutr 2018;108(2):363–70. doi: 10.1093/ajcn/nqy093 [published Online First: 2018/06/15]

70. Liu PH, Wu K, Ng K, et al. Association of Obesity With Risk of Early-Onset Colorectal Cancer Among Women. JAMA Oncol 2019;5(1):37–44. doi: 10.1001/jamaoncol.2018.4280 [published Online First: 2018/10/17]

71. Zhang X, Keum N, Wu K, et al. Calcium intake and colorectal cancer risk: Results from the nurses’ health study and health professionals follow-up study. Int J Cancer 2016;139(10):2232–42. doi: 10.1002/ijc.30293 [published Online First: 2016/07/29]

72. Nguyen LH, Liu PH, Zheng X, et al. Sedentary Behaviors, TV Viewing Time, and Risk of Young-Onset Colorectal Cancer. JNCI Cancer Spectr 2018;2(4):pky073. doi: 10.1093/jncics/pky073 [published Online First: 2019/02/12]

73. Patel SG, Ahnen DJ. Colorectal Cancer in the Young. Curr Gastroenterol Rep 2018;20(4):15. doi: 10.1007/s11894-018-0618-9 [published Online First: 2018/04/05]

