## Supplemental Table for "Sugar-sweetened beverage and sugar intake during adolescence and risk of colorectal cancer precursors: a large prospective U.S. cohort study": Suppl_Material.20201107.docx

**Supplementary Table 1.** Factor loadings for western and prudent dietary patterns from the high school food frequency questionnaire in the Nurses’ Health Study II

|  | **All foods** | |  | **All foods except soda^a^** | |  | **All foods except fruit juice^b^** | |
| --- | --- | --- | --- | --- | --- | --- | --- | --- |
| **Food Groups** | **Prudent** | **Western** |  | **Prudent** | **Western** |  | **Prudent** | **Western** |
| Other vegetables | 0.77 | . |  | 0.77 | . |  | 0.78 | . |
| Leafy green vegetables | 0.72 | . |  | 0.72 | . |  | 0.72 | . |
| Cruciferous vegetables | 0.68 | . |  | 0.68 | . |  | 0.68 | . |
| Yellow vegetables | 0.66 | . |  | 0.66 | . |  | 0.66 | . |
| Fruit | 0.65 | . |  | 0.65 | . |  | 0.64 | . |
| Tomatoes | 0.57 | . |  | 0.57 | . |  | 0.58 | . |
| Legumes | 0.52 | 0.16 |  | 0.51 | 0.18 |  | 0.53 | 0.17 |
| Salad dressing | 0.44 | 0.23 |  | 0.44 | . |  | 0.44 | 0.23 |
| Garlic | 0.43 | . |  | 0.43 | 0.24 |  | 0.44 | . |
| Fruit juice | 0.41 | . |  | 0.40 | . |  | – | – |
| Better quality grains | 0.39 | . |  | 0.38 | . |  | 0.38 | . |
| Fish | 0.37 | 0.17 |  | 0.37 | 0.18 |  | 0.37 | 0.18 |
| Poultry | 0.30 | 0.18 |  | 0.30 | 0.18 |  | 0.30 | 0.18 |
| Potato salad | 0.27 | 0.27 |  | 0.27 | 0.27 |  | 0.28 | 0.27 |
| Low-fat dairy | 0.26 | . |  | 0.26 | . |  | 0.25 | . |
| Organ meat | 0.19 | . |  | 0.19 | . |  | 0.20 | . |
| Tea | . | . |  | . | . |  | . | . |
| Diet soda | . | . |  | . | . |  | . | . |
| Coffee | . | . |  | . | . |  | . | . |
| Desserts and sweets | . | 0.62 |  | . | 0.62 |  | . | 0.62 |
| Condiments | 0.15 | 0.62 |  | . | 0.64 |  | 0.15 | 0.62 |
| Snack foods | 0.18 | 0.57 |  | 0.18 | 0.57 |  | 0.18 | 0.58 |
| Processed meat | . | 0.56 |  | . | 0.57 |  | . | 0.56 |
| Fries | . | 0.54 |  | . | 0.51 |  | . | 0.54 |
| Refined grains | . | 0.53 |  | . | 0.55 |  | . | 0.53 |
| Red meat | . | 0.52 |  | . | 0.53 |  | . | 0.52 |
| Mayonnaise | 0.19 | 0.44 |  | 0.17 | 0.45 |  | 0.19 | 0.44 |
| Nuts/peanut butter | 0.26 | 0.42 |  | 0.24 | 0.44 |  | 0.25 | 0.43 |
| High-fat dairy | . | 0.41 |  | . | 0.43 |  | . | 0.42 |
| Soda | -0.16 | 0.40 |  | – | – |  | -0.16 | 0.40 |
| Pizza | . | 0.36 |  | . | 0.34 |  | . | 0.36 |
| Potatos (mashed boiled) | . | 0.34 |  | . | 0.36 |  | . | 0.34 |
| Eggs | . | 0.32 |  | . | 0.34 |  | . | 0.31 |
| Margarine | . | 0.25 |  | . | 0.27 |  | . | 0.25 |
| Butter | . | 0.23 |  | . | 0.23 |  | . | 0.23 |
| Cream soup (Chowder) | 0.16 | 0.18 |  | . | 0.19 |  | 0.15 | 0.18 |
| Iced tea | . | 0.18 |  | . | 0.16 |  |  | 0.17 |
| Cereals | . | . |  | . | . |  | . | . |

Factor loadings are equivalent to Person correlation coefficients. Factor loadings <0.15 are not displayed for simplicity

**Supplementary Table 2.** Age-adjusted Spearman correlation coefficients of sweetened beverage and sugar intake during adolescence and adulthood

| **A. Correlation between adolescent intake of sweetened beverages and sugars** (n = 33106) | | | | | | | | | |
| --- | --- | --- | --- | --- | --- | --- | --- | --- | --- |
|  |  | **Adolescent intake** | | | | | | | |
|  |  | SSBs | ASBs | Fruit juice | Total fructose^a^ | Total glucose^a^ | Added sugar^a^ | Total sugar^a^ | Glycemic load |
| **Adolescent intake** | SSBs | 1 | -0.202 | 0.007 | 0.381 | 0.425 | 0.533 | 0.350 | 0.248 |
|  | ASBs |  | 1 | -0.014 | -0.061 | -0.071 | -0.074 | -0.126 | -0.085 |
|  | Fruit juice |  |  | 1 | 0.274 | 0.202 | -0.002 | 0.279 | 0.198 |
|  | Total fructose^a^ |  |  |  | 1 | 0.933 | 0.722 | 0.842 | 0.672 |
|  | Total glucose^a^ |  |  |  |  | 1 | 0.810 | 0.823 | 0.672 |
|  | Added sugar^a^ |  |  |  |  |  | 1 | 0.653 | 0.483 |
|  | Total sugar^a^ |  |  |  |  |  |  | 1 | 0.593 |
| **B. Correlation between adolescent and adult intake** (n = 52743) | | | | | | | | | |
|  |  | **Current (adult) intake** | | | | | | | |
|  |  | SSBs | ASBs | Fruit juice | Total fructose^a^ | Total glucose^a^ | Added sugar^a^ | Total sugar^a^ |  |
| **Adolescent intake** | SSBs | 0.253 |  |  |  |  |  |  |  |
|  | ASBs |  | 0.320 |  |  |  |  |  |  |
|  | Fruit juice |  |  | 0.374 |  |  |  |  |  |
|  | Total fructose^a^ |  |  |  | 0.265 |  |  |  |  |
|  | Total glucose^a^ |  |  |  |  | 0.254 |  |  |  |
|  | Added sugar^a^ |  |  |  |  |  | 0.253 |  |  |
|  | Total sugar^a^ |  |  |  |  |  |  | 0.253 |  |

SSBs, sugar-sweetende beverages; ASBs, artificially-sweetened beverages

^a^From mono- and disaccharide sugars

**Supplementary Table 3.** Odds ratios and 95% confidence intervals of CRC precursor according to glucose, added sugar, and total sugar intake during adolescence in the Nurses’ Health Study II, 1998-2015

|  | **Glucose intake during adolescence, % of calorie** | | | | | | **Per 5% of calorie/d increase** |
| --- | --- | --- | --- | --- | --- | --- | --- |
|  | **Q1** | **Q2** | **Q3** | **Q4** | **Q5** | ***P*_trend_** |  |
| **Total adenoma** |  |  |  |  |  |  |  |
| *N*_cases_/*N*_controls_ | 564/9984 | 598/9951 | 574/9974 | 593/9957 | 580/9968 |  |  |
| Model 1^a^ | 1 (ref) | 1.07 (0.95-1.20) | 1.03 (0.92-1.16) | 1.07 (0.95-1.20) | 1.05 (0.93-1.19) | 0.466 | 1.04 (0.94-1.14) |
| Model 2^b^ | 1 (ref) | 1.07 (0.95-1.21) | 1.04 (0.92-1.18) | 1.08 (0.96-1.22) | 1.08 (0.95-1.21) | 0.267 | 1.05 (0.96-1.16) |
| Model 3^c^ | 1 (ref) | 1.12 (0.99-1.26) | 1.11 (0.98-1.26) | 1.18 (1.03-1.34) | 1.18 (1.03-1.36) | 0.023 | 1.14 (1.02-1.27) |
| Proximal adenoma |  |  |  |  |  |  |  |
| *N*_cases_/*N*_controls_ | 311/9806 | 313/9771 | 323/9793 | 314/9780 | 287/9802 |  |  |
| Model 3 | 1 (ref) | 1.07 (0.91-1.26) | 1.16 (0.97-1.38) | 1.15 (0.96-1.39) | 1.09 (0.90-1.32) | 0.347 | 1.08 (0.92-1.26) |
| Distal adenoma |  |  |  |  |  |  |  |
| *N*_cases_/*N*_controls_ | 233/9806 | 241/9771 | 226/9793 | 248/9780 | 257/9802 |  |  |
| Model 3 | 1 (ref) | 1.09 (0.90-1.32) | 1.06 (0.86-1.30) | 1.20 (0.98-1.47) | 1.28 (1.03-1.58) | 0.020 | 1.23 (1.03-1.46) |
| Rectal adenoma |  |  |  |  |  |  |  |
| *N*_cases_/*N*_controls_ | 79/9806 | 100/9771 | 84/9793 | 91/9780 | 104/9802 |  |  |
| Model 3 | 1 (ref) | 1.35 (0.99-1.85) | 1.18 (0.85-1.63) | 1.32 (0.94-1.84) | 1.55 (1.09-2.19) | 0.030 | 1.35 (1.03-1.78) |
| Low-risk adenoma^d^ |  |  |  |  |  |  |  |
| *N*_cases_/*N*_controls_ | 291/9806 | 290/9771 | 282/9793 | 256/9780 | 265/9802 |  |  |
| Model 3 | 1 (ref) | 1.03 (0.87-1.23) | 1.03 (0.86-1.24) | 0.95 (0.78-1.15) | 1.01 (0.83-1.24) | 0.863 | 0.99 (0.84-1.16) |
| High-risk adenoma^d^ |  |  |  |  |  |  |  |
| *N*_cases_/*N*_controls_ | 193/9806 | 215/9771 | 207/9793 | 242/9780 | 225/9802 |  |  |
| Model 3 | 1 (ref) | 1.19 (0.97-1.46) | 1.21 (0.97-1.50) | 1.45 (1.17-1.81) | 1.38 (1.09-1.74) | 0.003 | 1.32 (1.10-1.58) |
| **Total serrated lesions**^e^ |  |  |  |  |  |  |  |
| *N*_cases_/*N*_controls_ | 457/9980 | 469/9965 | 471/9952 | 495/9928 | 463/9981 |  |  |
| Model 3 | 1 (ref) | 1.03 (0.89-1.19) | 1.06 (0.91-1.22) | 1.13 (0.97-1.31) | 1.06 (0.90-1.24) | 0.337 | 1.06 (0.94-1.21) |
| Small serrated lesion^f^ |  |  |  |  |  |  |  |
| *N*_cases_/*N*_controls_ | 382/9980 | 406/9965 | 408/9952 | 426/9928 | 393/9981 |  |  |
| Model 3 | 1 (ref) | 1.07 (0.92-1.25) | 1.10 (0.94-1.29) | 1.18 (1.00-1.38) | 1.09 (0.92-1.30) | 0.235 | 1.09 (0.95-1.24) |
| Large serrated lesion^f^ |  |  |  |  |  |  |  |
| *N*_cases_/*N*_controls_ | 49/9980 | 35/9965 | 37/9952 | 42/9928 | 33/9981 |  |  |
| Model 3 | 1 (ref) | 0.69 (0.44-1.09) | 0.74 (0.46-1.18) | 0.81 (0.52-1.28) | 0.64 (0.40-1.01) | 0.142 | 0.75 (0.52-1.10) |
| **Subtype of CRC precursor** |  |  |  |  |  |  |  |
| Adenoma only |  |  |  |  |  |  |  |
| *N*_cases_/*N*_controls_ | 470/9621 | 509/9571 | 453/9624 | 481/9574 | 476/9609 |  |  |
| Model 3 | 1 (ref) | 1.15 (1.01-1.32) | 1.06 (0.92-1.22) | 1.15 (1.00-1.34) | 1.17 (1.00-1.36) | 0.088 | 1.11 (0.98-1.26) |
| Serrated lesion only |  |  |  |  |  |  |  |
| *N*_cases_/*N*_controls_ | 363/9621 | 380/9571 | 350/9624 | 383/9574 | 359/9609 |  |  |
| Model 3 | 1 (ref) | 1.06 (0.91-1.23) | 0.97 (0.83-1.15) | 1.09 (0.92-1.29) | 1.02 (0.86-1.22) | 0.770 | 1.02 (0.89-1.17) |
| Both adenoma and serrated lesion |  |  |  |  |  |  |  |
| *N*_cases_/*N*_controls_ | 94/9621 | 89/9571 | 121/9624 | 112/9574 | 104/9609 |  |  |
| Model 3 | 1 (ref) | 0.97 (0.72-1.31) | 1.36 (1.02-1.83) | 1.32 (0.97-1.79) | 1.27 (0.92-1.77) | 0.061 | 1.27 (0.99-1.64) |

**Supplementary Table 3. (cont.)**

|  | **Added sugar intake during adolescence, % of calorie** | | | | | | **Per 10% of calorie/d increase** |
| --- | --- | --- | --- | --- | --- | --- | --- |
|  | **Q1** | **Q2** | **Q3** | **Q4** | **Q5** | ***P*_trend_** |  |
| **Total adenoma** |  |  |  |  |  |  |  |
| *N*_cases_/*N*_controls_ | 568/9979 | 580/9970 | 550/9999 | 579/9970 | 632/9916 |  |  |
| Model 1^a^ | 1 (ref) | 1.02 (0.91-1.15) | 0.97 (0.86-1.10) | 1.03 (0.92-1.16) | 1.14 (1.01-1.28) | 0.027 | 1.12 (1.01-1.23) |
| Model 2^b^ | 1 (ref) | 1.02 (0.90-1.15) | 0.96 (0.85-1.09) | 1.02 (0.90-1.15) | 1.13 (1.00-1.27) | 0.039 | 1.11 (1.01-1.22) |
| Model 3^c^ | 1 (ref) | 1.01 (0.90-1.15) | 0.97 (0.85-1.10) | 1.04 (0.91-1.19) | 1.16 (1.01-1.33) | 0.025 | 1.14 (1.02-1.28) |
| Proximal adenoma |  |  |  |  |  |  |  |
| *N*_cases_/*N*_controls_ | 306/9794 | 309/9801 | 301/9821 | 307/9801 | 325/9735 |  |  |
| Model 3 | 1 (ref) | 0.99 (0.84-1.17) | 0.98 (0.83-1.17) | 1.03 (0.86-1.23) | 1.10 (0.91-1.33) | 0.250 | 1.10 (0.94-1.29) |
| Distal adenoma |  |  |  |  |  |  |  |
| *N*_cases_/*N*_controls_ | 241/9794 | 235/9801 | 221/9821 | 238/9801 | 270/9735 |  |  |
| Model 3 | 1 (ref) | 0.97 (0.81-1.18) | 0.92 (0.75-1.12) | 1.02 (0.83-1.25) | 1.20 (0.97-1.48) | 0.060 | 1.19 (0.99-1.42) |
| Rectal adenoma |  |  |  |  |  |  |  |
| *N*_cases_/*N*_controls_ | 84/9794 | 87/9801 | 85/9821 | 93/9801 | 109/9735 |  |  |
| Model 3 | 1 (ref) | 1.06 (0.77-1.45) | 1.06 (0.76-1.46) | 1.18 (0.85-1.65) | 1.35 (0.96-1.91) | 0.062 | 1.31 (0.99-1.74) |
| Low-risk adenoma^d^ |  |  |  |  |  |  |  |
| *N*_cases_/*N*_controls_ | 278/9794 | 282/9801 | 266/9821 | 268/9801 | 290/9735 |  |  |
| Model 3 | 1 (ref) | 0.99 (0.83-1.18) | 0.94 (0.78-1.13) | 0.96 (0.80-1.16) | 1.07 (0.88-1.30) | 0.519 | 1.06 (0.89-1.25) |
| High-risk adenoma^d^ |  |  |  |  |  |  |  |
| *N*_cases_/*N*_controls_ | 206/9794 | 210/9801 | 194/9821 | 228/9801 | 244/9735 |  |  |
| Model 3 | 1 (ref) | 1.01 (0.82-1.23) | 0.94 (0.76-1.16) | 1.13 (0.91-1.41) | 1.21 (0.97-1.52) | 0.045 | 1.21 (1.00-1.47) |
| **Total serrated lesions**^e^ |  |  |  |  |  |  |  |
| *N*_cases_/*N*_controls_ | 444/10000 | 448/9982 | 454/9971 | 524/9899 | 485/9954 |  |  |
| Model 3 | 1 (ref) | 0.99 (0.86-1.14) | 1.00 (0.87-1.16) | 1.17 (1.01-1.35) | 1.08 (0.92-1.27) | 0.110 | 1.11 (0.98-1.26) |
| Small serrated lesion^f^ |  |  |  |  |  |  |  |
| *N*_cases_/*N*_controls_ | 373/10000 | 387/9982 | 394/9971 | 450/9899 | 411/9954 |  |  |
| Model 3 | 1 (ref) | 1.02 (0.87-1.19) | 1.05 (0.90-1.22) | 1.21 (1.04-1.42) | 1.11 (0.94-1.32) | 0.079 | 1.13 (0.99-1.30) |
| Large serrated lesion^f^ |  |  |  |  |  |  |  |
| *N*_cases_/*N*_controls_ | 41/10000 | 39/9982 | 33/9971 | 43/9899 | 40/9954 |  |  |
| Model 3 | 1 (ref) | 0.90 (0.57-1.42) | 0.75 (0.46-1.23) | 0.94 (0.60-1.48) | 0.87 (0.53-1.41) | 0.702 | 0.92 (0.61-1.39) |
| **Subtype of CRC precursor** |  |  |  |  |  |  |  |
| Adenoma only |  |  |  |  |  |  |  |
| *N*_cases_/*N*_controls_ | 473/9630 | 481/9621 | 449/9646 | 473/9552 | 513/9550 |  |  |
| Model 3 | 1 (ref) | 1.02 (0.89-1.17) | 0.96 (0.83-1.11) | 1.04 (0.90-1.21) | 1.14 (0.98-1.33) | 0.067 | 1.13 (0.99-1.28) |
| Serrated lesion only |  |  |  |  |  |  |  |
| *N*_cases_/*N*_controls_ | 349/9630 | 349/9621 | 353/9646 | 418/9552 | 366/9550 |  |  |
| Model 3 | 1 (ref) | 0.98 (0.84-1.15) | 0.99 (0.85-1.17) | 1.19 (1.01-1.40) | 1.05 (0.88-1.25) | 0.235 | 1.09 (0.95-1.26) |
| Both adenoma and serrated lesion |  |  |  |  |  |  |  |
| *N*_cases_/*N*_controls_ | 95/9630 | 99/9621 | 101/9646 | 106/9552 | 119/9550 |  |  |
| Model 3 | 1 (ref) | 0.98 (0.73-1.31) | 1.01 (0.75-1.36) | 1.07 (0.79-1.44) | 1.25 (0.92-1.71) | 0.109 | 1.24 (0.95-1.61) |

**Supplementary Table 3. (cont.)**

|  | **Total sugar intake during adolescence, % of calorie** | | | | | | **Per 10% of calorie/d increase** |
| --- | --- | --- | --- | --- | --- | --- | --- |
|  | **Q1** | **Q2** | **Q3** | **Q4** | **Q5** | ***P*_trend_** |  |
| **Total adenoma** |  |  |  |  |  |  |  |
| *N*_cases_/*N*_controls_ | 557/9990 | 563/9987 | 620/9928 | 568/9982 | 601/9947 |  |  |
| Model 1^a^ | 1 (ref) | 1.01(0.89-1.14) | 1.12(1.00-1.27) | 1.04(0.92-1.17) | 1.10(0.97-1.24) | 0.117 | 1.08(0.98-1.19) |
| Model 2^b^ | 1 (ref) | 1.02(0.90-1.15) | 1.15(1.02-1.29) | 1.06(0.94-1.20) | 1.14(1.01-1.28) | 0.030 | 1.11(1.01-1.23) |
| Model 3^c^ | 1 (ref) | 1.02(0.90-1.16) | 1.17(1.03-1.33) | 1.10(0.96-1.25) | 1.20(1.05-1.38) | 0.005 | 1.18(1.05-1.31) |
| Proximal adenoma |  |  |  |  |  |  |  |
| *N*_cases_/*N*_controls_ | 301/9824 | 312/9787 | 323/9760 | 298/9817 | 314/9764 |  |  |
| Model 3 | 1 (ref) | 1.05(0.89-1.24) | 1.13(0.95-1.34) | 1.07(0.89-1.28) | 1.17(0.97-1.41) | 0.105 | 1.14(0.97-1.32) |
| Distal adenoma |  |  |  |  |  |  |  |
| *N*_cases_/*N*_controls_ | 229/9824 | 224/9787 | 241/9760 | 252/9817 | 259/9764 |  |  |
| Model 3 | 1 (ref) | 1.00(0.82-1.22) | 1.13(0.93-1.37) | 1.21(0.99-1.48) | 1.29(1.05-1.59) | 0.005 | 1.29(1.08-1.53) |
| Rectal adenoma |  |  |  |  |  |  |  |
| *N*_cases_/*N*_controls_ | 81/9824 | 81/9787 | 112/9760 | 83/9817 | 101/9764 |  |  |
| Model 3 | 1 (ref) | 1.05(0.77-1.44) | 1.57(1.16-2.14) | 1.19(0.86-1.64) | 1.53(1.11-2.12) | 0.009 | 1.42(1.09-1.85) |
| Low-risk adenoma^d^ |  |  |  |  |  |  |  |
| *N*_cases_/*N*_controls_ | 272/9824 | 291/9787 | 274/9760 | 277/9817 | 270/9764 |  |  |
| Model 3 | 1 (ref) | 1.07(0.90-1.28) | 1.04(0.86-1.25) | 1.07(0.88-1.29) | 1.09(0.89-1.32) | 0.484 | 1.06(0.90-1.25) |
| High-risk adenoma^d^ |  |  |  |  |  |  |  |
| *N*_cases_/*N*_controls_ | 198/9824 | 202/9787 | 237/9760 | 208/9817 | 237/9764 |  |  |
| Model 3 | 1 (ref) | 1.07(0.87-1.31) | 1.32(1.08-1.62) | 1.20(0.96-1.48) | 1.41(1.14-1.76) | 0.001 | 1.34(1.12-1.61) |
| **Total serrated lesions**^e^ |  |  |  |  |  |  |  |
| *N*_cases_/*N*_controls_ | 462/9986 | 447/9981 | 505/9919 | 469/9956 | 472/9964 |  |  |
| Model 3 | 1 (ref) | 0.98(0.85-1.12) | 1.17(1.02-1.35) | 1.10(0.95-1.27) | 1.12(0.97-1.31) | 0.052 | 1.13(1.00-1.28) |
| Small serrated lesion^f^ |  |  |  |  |  |  |  |
| *N*_cases_/*N*_controls_ | 391/9986 | 381/9981 | 433/9919 | 410/9956 | 400/9964 |  |  |
| Model 3 | 1 (ref) | 0.98(0.84-1.14) | 1.19(1.02-1.39) | 1.15(0.98-1.34) | 1.14(0.97-1.34) | 0.040 | 1.15(1.01-1.31) |
| Large serrated lesion^f^ |  |  |  |  |  |  |  |
| *N*_cases_/*N*_controls_ | 43/9986 | 45/9981 | 44/9919 | 28/9956 | 36/9964 |  |  |
| Model 3 | 1 (ref) | 1.07(0.69-1.67) | 1.09(0.69-1.74) | 0.68(0.41-1.13) | 0.91(0.57-1.45) | 0.318 | 0.82(0.56-1.21) |
| **Subtype of CRC precursor** |  |  |  |  |  |  |  |
| Adenoma only |  |  |  |  |  |  |  |
| *N*_cases_/*N*_controls_ | 461/9624 | 465/9638 | 514/9529 | 457/9624 | 492/9584 |  |  |
| Model 3 | 1 (ref) | 1.02(0.89-1.17) | 1.18(1.02-1.35) | 1.06(0.91-1.22) | 1.18(1.02-1.37) | 0.028 | 1.15(1.02-1.30) |
| Serrated lesion only |  |  |  |  |  |  |  |
| *N*_cases_/*N*_controls_ | 366/9624 | 349/9638 | 399/9529 | 358/9624 | 363/9584 |  |  |
| Model 3 | 1 (ref) | 0.96(0.82-1.12) | 1.17(1.00-1.36) | 1.05(0.89-1.23) | 1.08(0.92-1.28) | 0.221 | 1.09(0.95-1.25) |
| Both adenoma and serrated lesion |  |  |  |  |  |  |  |
| *N*_cases_/*N*_controls_ | 96/9624 | 98/9638 | 106/9529 | 111/9624 | 109/9584 |  |  |
| Model 3 | 1 (ref) | 1.02(0.76-1.36) | 1.21(0.90-1.62) | 1.30(0.96-1.76) | 1.33(0.97-1.81) | 0.030 | 1.33(1.03-1.72) |

Q, quintile; N, number of endoscopies

^a^Adjusted for age, time period of endoscopy, number of endoscopies during the study period (continuous), time since most recent endoscopy (continuous), and reason for current endoscopy (screening/symptoms)

^b^Additionally adjusted for family history of colorectal cancer (yes/no), menopausal status/menopausal hormone use (premenopausal, postmenopausal with never, past, or current hormone therapy), current aspirin use ≥2 times/wk (yes/no), history of type 2 diabetes (yes/no), adult height (continuous), BMI at age 18 y (<18.5, 18.5–<20, 20–<22.5, 22.5–<25, ≥25 kg/m^2^), current BMI (<22.5, 22.5–<25, 25–<27.5, 27.5–<30, ≥30 kg/m^2^), smoking status at 19 y (never, 0≤2.5, >2.5 pack-years), current smoking status (never smoker, past smoker <30 pack-years, past smoker ≥30 pack-years, current smoker <30 pack-years, current smoker ≥30 pack-years), alcohol intake at 18–22 y (<0.1, 0.1–4.9, 5–14.9, ≥15 g/d), current alcohol intake (none, 0.1–4.9, 5–9.9, 10–14.9, ≥15 g/d), physical activity during grades 9–12 (quintile), and current physical activity (<21, 21–<30, 30–<39, 39–<54, ≥54 MET hours/wk)

^c^Additionally adjusted for adolescent and current (adult) dietary intake (total calorie, total calcium, vitamin D, total folate, total fiber, fruits, vegetables, and dairy; quintile), current total red meat intake (quintile), a western dietary pattern score during adolescence (quintile), and corresponding current sugar intake (quintiles) in Model 2

^d^Low risk: small(<1cm), tubular, and single adenoma; high risk: large (≥1 cm) or villous adenoma or more than two adenoma

^e^Included hyperplastic polyp, sessile serrated adenoma/polyp, and traditional serrated adenoma

^f^Small, <1cm; large, ≥1 cm

**Supplementary Table 4.** Odds ratios and 95% confidence intervals of CRC precursor according to artificially-sweetened beverage and fruit juice intake during adolescence in the Nurses’ Health Study II, 1998-2015

|  | **Artificially-sweetened beverage intake during adolescence, servings** | | | | | **Per 1 serving/d increase** |
| --- | --- | --- | --- | --- | --- | --- |
|  | **<1/week** | **1-6/week** | **1/day** | **≥2/day** | ***P*_trend_** |  |
| **Total adenoma** |  |  |  |  |  |  |
| *N*_cases_/*N*_controls_ | 2017/34000 | 582/10361 | 159/2860 | 151/2613 |  |  |
| Model 1^a^ | 1 (ref) | 0.96(0.88-1.06) | 0.97(0.82-1.15) | 1.01(0.85-1.20) | 0.952 | 1.00(0.93-1.07) |
| Model 2^b^ | 1 (ref) | 0.94(0.85-1.03) | 0.92(0.78-1.09) | 0.96(0.80-1.14) | 0.397 | 0.97(0.91-1.04) |
| Model 3^c^ | 1 (ref) | 0.95(0.85-1.05) | 0.93(0.78-1.11) | 0.94(0.79-1.13) | 0.402 | 0.97(0.90-1.04) |
| Proximal adenoma |  |  |  |  |  |  |
| *N*_cases_/*N*_controls_ | 1058/33373 | 323/10191 | 87/2816 | 80/2572 |  |  |
| Model 3 | 1 (ref) | 1.01(0.88-1.16) | 0.97(0.77-1.23) | 0.96(0.75-1.24) | 0.749 | 0.98(0.89-1.08) |
| Distal adenoma |  |  |  |  |  |  |
| *N*_cases_/*N*_controls_ | 837/33373 | 244/10191 | 59/2816 | 65/2572 |  |  |
| Model 3 | 1 (ref) | 0.93(0.80-1.09) | 0.80(0.61-1.06) | 0.93(0.71-1.23) | 0.370 | 0.95(0.85-1.06) |
| Rectal adenoma |  |  |  |  |  |  |
| *N*_cases_/*N*_controls_ | 327/33373 | 80/10191 | 32/2816 | 19/2572 |  |  |
| Model 3 | 1 (ref) | 0.82(0.63-1.07) | 1.13(0.76-1.67) | 0.69(0.42-1.13) | 0.217 | 0.89(0.75-1.07) |
| Low-risk adenoma^d^ |  |  |  |  |  |  |
| *N*_cases_/*N*_controls_ | 959/33373 | 293/10191 | 65/2816 | 67/2572 |  |  |
| Model 3 | 1 (ref) | 0.99(0.86-1.15) | 0.80(0.61-1.04) | 0.90(0.69-1.17) | 0.230 | 0.94(0.85-1.04) |
| High-risk adenoma^d^ |  |  |  |  |  |  |
| *N*_cases_/*N*_controls_ | 736/33373 | 212/10191 | 77/2816 | 57/2572 |  |  |
| Model 3 | 1 (ref) | 0.97(0.81-1.15) | 1.24(0.96-1.60) | 0.97(0.73-1.31) | 0.755 | 1.02(0.91-1.13) |
| **Total serrated lesions**^e^ |  |  |  |  |  |  |
| *N*_cases_/*N*_controls_ | 1599/34022 | 504/10303 | 130/2859 | 122/2622 |  |  |
| Model 3 | 1 (ref) | 1.03(0.92-1.15) | 0.93(0.77-1.14) | 0.96(0.78-1.18) | 0.588 | 0.98(0.90-1.06) |
| Small serrated lesion^f^ |  |  |  |  |  |  |
| *N*_cases_/*N*_controls_ | 1362/34022 | 436/10303 | 112/2859 | 105/2622 |  |  |
| Model 3 | 1 (ref) | 1.05(0.93-1.18) | 0.96(0.78-1.18) | 0.98(0.78-1.22) | 0.786 | 0.99(0.91-1.07) |
| Large serrated lesion^f^ |  |  |  |  |  |  |
| *N*_cases_/*N*_controls_ | 134/34022 | 45/10303 | 8/2859 | 9/2622 |  |  |
| Model 3 | 1 (ref) | 1.05(0.72-1.53) | 0.63(0.30-1.30) | 0.78(0.38-1.59) | 0.340 | 0.87(0.66-1.15) |
| **Subtype of CRC precursor** |  |  |  |  |  |  |
| Adenoma only |  |  |  |  |  |  |
| *N*_cases_/*N*_controls_ | 1669/32749 | 470/9969 | 132/2757 | 118/2524 |  |  |
| Model 3 | 1 (ref) | 0.93(0.83-1.04) | 0.94(0.78-1.14) | 0.89(0.73-1.09) | 0.224 | 0.95(0.88-1.03) |
| Serrated lesion only |  |  |  |  |  |  |
| *N*_cases_/*N*_controls_ | 1251/32749 | 392/9969 | 103/2757 | 89/2524 |  |  |
| Model 3 | 1 (ref) | 1.01(0.89-1.14) | 0.95(0.77-1.18) | 0.90(0.71-1.14) | 0.342 | 0.96(0.88-1.05) |
| Both adenoma and serrated lesion |  |  |  |  |  |  |
| *N*_cases_/*N*_controls_ | 348/32749 | 112/9969 | 27/2757 | 33/2524 |  |  |
| Model 3 | 1 (ref) | 1.03(0.82-1.30) | 0.86(0.57-1.30) | 1.16(0.78-1.70) | 0.639 | 1.04(0.89-1.21) |

**Supplementary Table 4. (cont.)**

|  | **Fruit juice intake during adolescence, servings** | | | | | **Per 1 serving/d increase** |
| --- | --- | --- | --- | --- | --- | --- |
|  | **<1/week** | **1-6/week** | **1/day** | **≥2/day** | ***P*_trend_** |  |
| **Total adenoma** |  |  |  |  |  |  |
| *N*_cases_/*N*_controls_ | 559/9392 | 1431/24656 | 805/13706 | 114/2080 |  |  |
| Model 1^a^ | 1 (ref) | 0.98(0.89-1.09) | 1.00(0.89-1.11) | 0.95(0.77-1.17) | 0.768 | 0.99(0.92-1.06) |
| Model 2^b^ | 1 (ref) | 1.00(0.90-1.11) | 1.03(0.92-1.16) | 0.99(0.80-1.22) | 0.715 | 1.01(0.94-1.09) |
| Model 3^c^ | 1 (ref) | 1.00(0.90-1.12) | 1.08(0.94-1.24) | 1.10(0.87-1.40) | 0.206 | 1.06(0.97-1.16) |
| Proximal adenoma |  |  |  |  |  |  |
| *N*_cases_/*N*_controls_ | 280/9235 | 795/24216 | 418/13463 | 55/2038 |  |  |
| Model 3 | 1 (ref) | 1.13(0.97-1.31) | 1.13(0.93-1.38) | 1.10(0.78-1.54) | 0.536 | 1.04(0.92-1.17) |
| Distal adenoma |  |  |  |  |  |  |
| *N*_cases_/*N*_controls_ | 231/9235 | 575/24216 | 345/13463 | 54/2038 |  |  |
| Model 3 | 1 (ref) | 0.98(0.82-1.16) | 1.10(0.89-1.36) | 1.23(0.86-1.75) | 0.137 | 1.11(0.97-1.27) |
| Rectal adenoma |  |  |  |  |  |  |
| *N*_cases_/*N*_controls_ | 110/9235 | 215/24216 | 119/13463 | 14/2038 |  |  |
| Model 3 | 1 (ref) | 0.78(0.60-1.00) | 0.89(0.65-1.21) | 0.73(0.40-1.34) | 0.550 | 0.94(0.75-1.17) |
| Low-risk adenoma^d^ |  |  |  |  |  |  |
| *N*_cases_/*N*_controls_ | 277/9235 | 687/24216 | 368/13463 | 52/2038 |  |  |
| Model 3 | 1 (ref) | 0.95(0.81-1.11) | 0.97(0.79-1.18) | 0.99(0.70-1.41) | 0.988 | 1.00(0.88-1.14) |
| High-risk adenoma^d^ |  |  |  |  |  |  |
| *N*_cases_/*N*_controls_ | 208/9235 | 529/24216 | 302/13463 | 43/2038 |  |  |
| Model 3 | 1 (ref) | 1.02(0.85-1.21) | 1.14(0.91-1.42) | 1.17(0.80-1.71) | 0.221 | 1.09(0.95-1.26) |
| **Total serrated lesions**^e^ |  |  |  |  |  |  |
| *N*_cases_/*N*_controls_ | 498/9337 | 1150/24649 | 620/13741 | 87/2079 |  |  |
| Model 3 | 1 (ref) | 0.84(0.74-0.95) | 0.83(0.71-0.97) | 0.81(0.62-1.06) | 0.138 | 0.92(0.83-1.03) |
| Small serrated lesion^f^ |  |  |  |  |  |  |
| *N*_cases_/*N*_controls_ | 429/9337 | 977/24649 | 539/13741 | 70/2079 |  |  |
| Model 3 | 1 (ref) | 0.83(0.73-0.94) | 0.85(0.72-1.00) | 0.77(0.58-1.04) | 0.154 | 0.92(0.82-1.03) |
| Large serrated lesion^f^ |  |  |  |  |  |  |
| *N*_cases_/*N*_controls_ | 43/9337 | 100/24649 | 45/13741 | 8/2079 |  |  |
| Model 3 | 1 (ref) | 0.93(0.63-1.36) | 0.79(0.47-1.33) | 1.02(0.43-2.43) | 0.755 | 0.94(0.65-1.37) |
| **Subtype of CRC precursor** |  |  |  |  |  |  |
| Adenoma only |  |  |  |  |  |  |
| *N*_cases_/*N*_controls_ | 459/8994 | 1178/23759 | 659/13232 | 93/2014 |  |  |
| Model 3 | 1 (ref) | 1.00(0.89-1.13) | 1.08(0.93-1.25) | 1.08(0.83-1.41) | 0.311 | 1.05(0.95-1.16) |
| Serrated lesion only |  |  |  |  |  |  |
| *N*_cases_/*N*_controls_ | 398/8994 | 897/23759 | 474/13232 | 66/2014 |  |  |
| Model 3 | 1 (ref) | 0.82(0.72-0.93) | 0.79(0.66-0.94) | 0.74(0.55-0.99) | 0.042 | 0.88(0.79-1.00) |
| Both adenoma and serrated lesion |  |  |  |  |  |  |
| *N*_cases_/*N*_controls_ | 100/8994 | 253/23759 | 146/13232 | 21/2014 |  |  |
| Model 3 | 1 (ref) | 0.94(0.72-1.21) | 1.02(0.74-1.42) | 1.10(0.64-1.89) | 0.588 | 1.06(0.86-1.30) |

^a^Adjusted for age, time period of endoscopy, number of endoscopies during the study period (continuous), time since most recent endoscopy (continuous), and reason for current endoscopy (screening/symptoms)

^b^Additionally adjusted for family history of colorectal cancer, menopausal status/menopausal hormone use, current aspirin use ≥2 times/wk, history of type 2 diabetes, adult height (continuous), BMI at age 18 y, current BMI, smoking status at 19 y, current smoking status, alcohol intake at 18–22 y, current alcohol intake, physical activity during grades 9–12, current physical activity

^c^Additionally adjusted for adolescent and current (adult) dietary intake (total calorie, total calcium, vitamin D, total folate, total fiber, fruit, vegetables, and dairy), current total red meat intake (quintile), a western dietary pattern score during adolescence (quintile), and corresponding current beverage intake in Model 2

**Supplementary Table 5.** Odds ratios and 95% confidence intervals of colorectal adenoma according to sugar-sweetened beverage (SSB) and total fructose intake during adolescence after additional adjustment for dietary factors

|  | **Sugar-sweetened beverage intake during adolescence, servings** | | | | | **Per 1 serving/d increase** |
| --- | --- | --- | --- | --- | --- | --- |
|  | **<1/week** | **1-6/week** | **1/day** | **≥2/day** | ***P*_trend_** |  |
| **Model 3^a^** |  |  |  |  |  |  |
| Total adenoma | 1 (ref) | 0.92 (0.84-1.00) | 1.20 (1.03-1.39) | 1.21 (1.00-1.47) | 0.012 | 1.10 (1.02-1.19) |
| Proximal adenoma | 1 (ref) | 0.91 (0.81-1.03) | 1.25 (1.03-1.53) | 1.27 (0.97-1.66) | 0.023 | 1.13 (1.02-1.26) |
| Rectal adenoma | 1 (ref) | 0.85 (0.68-1.06) | 1.45 (1.03-2.05) | 1.62 (1.03-2.53) | 0.008 | 1.28 (1.07-1.53) |
| High-risk adenoma | 1 (ref) | 0.89 (0.77-1.03) | 1.25 (0.99-1.58) | 1.21 (0.88-1.65) | 0.085 | 1.11 (0.99-1.26) |
| **Model 3 + red meat intake**^b^ |  |  |  |  |  |  |
| Total adenoma | 1 (ref) | 0.92 (0.84-1.00) | 1.19 (1.03-1.38) | 1.21 (0.99-1.47) | 0.013 | 1.10 (1.02-1.19) |
| Proximal adenoma | 1 (ref) | 0.91 (0.81-1.03) | 1.26 (1.03-1.53) | 1.27 (0.97-1.67) | 0.022 | 1.13 (1.02-1.26) |
| Rectal adenoma | 1 (ref) | 0.85 (0.68-1.06) | 1.46 (1.04-2.05) | 1.63 (1.04-2.54) | 0.007 | 1.28 (1.07-1.53) |
| High-risk adenoma | 1 (ref) | 0.89 (0.77-1.03) | 1.26 (1.00-1.58) | 1.21 (0.88-1.65) | 0.083 | 1.12 (0.99-1.26) |
| **Model 3 + prudent dietary pattern**^b^ **(in place of western dietary pattern)** | | | | | | |
| Total adenoma | 1 (ref) | 0.92 (0.84-1.00) | 1.18 (1.02-1.37) | 1.19 (0.98-1.44) | 0.017 | 1.10 (1.02-1.18) |
| Proximal adenoma | 1 (ref) | 0.91 (0.81-1.02) | 1.25 (1.02-1.52) | 1.26 (0.97-1.64) | 0.022 | 1.13 (1.02-1.25) |
| Rectal adenoma | 1 (ref) | 0.83 (0.67-1.04) | 1.40 (1.00-1.97) | 1.53 (0.99-2.37) | 0.013 | 1.25 (1.05-1.49) |
| High-risk adenoma | 1 (ref) | 0.89 (0.77-1.02) | 1.25 (0.99-1.57) | 1.21 (0.89-1.64) | 0.078 | 1.12 (0.99-1.26) |
| **Model 3 + prudent dietary pattern**^b^ **(in addition to western dietary pattern)** | | | | | | |
| Total adenoma | 1 (ref) | 0.91 (0.84-1.00) | 1.18 (1.02-1.37) | 1.18 (0.97-1.44) | 0.024 | 1.09 (1.01-1.18) |
| Proximal adenoma | 1 (ref) | 0.90 (0.80-1.02) | 1.23 (1.01-1.51) | 1.24 (0.94-1.62) | 0.037 | 1.12 (1.01-1.25) |
| Rectal adenoma | 1 (ref) | 0.83 (0.67-1.04) | 1.40 (0.99-1.97) | 1.53 (0.98-2.38) | 0.017 | 1.25 (1.04-1.49) |
| High-risk adenoma | 1 (ref) | 0.89 (0.77-1.02) | 1.24 (0.99-1.57) | 1.19 (0.87-1.63) | 0.098 | 1.11 (0.98-1.26) |
| **Model 3 + artificially-sweetened beverage + fruit juice intake**^c^ | | | | | | |
| Total adenoma | 1 (ref) | 0.91 (0.83-1.00) | 1.18 (1.02-1.37) | 1.20 (0.98-1.46) | 0.018 | 1.10 (1.02-1.19) |
| Proximal adenoma | 1 (ref) | 0.90 (0.80-1.02) | 1.24 (1.02-1.52) | 1.26 (0.96-1.65) | 0.026 | 1.13 (1.01-1.26) |
| Rectal adenoma | 1 (ref) | 0.85 (0.68-1.07) | 1.43 (1.01-2.02) | 1.60 (1.02-2.51) | 0.009 | 1.27 (1.06-1.52) |
| High-risk adenoma | 1 (ref) | 0.90 (0.77-1.04) | 1.25 (0.99-1.58) | 1.23 (0.90-1.68) | 0.066 | 1.12 (0.99-1.27) |
| **Model 3 + glycemic index**^b^ | | | | | | |
| Total adenoma | 1 (ref) | 0.91 (0.83-0.99) | 1.15 (0.99-1.34) | 1.13 (0.92-1.39) | 0.078 | 1.08 (0.99-1.17) |
| Proximal adenoma | 1 (ref) | 0.90 (0.79-1.01) | 1.20 (0.98-1.47) | 1.16 (0.88-1.53) | 0.120 | 1.09 (0.98-1.22) |
| Rectal adenoma | 1 (ref) | 0.84 (0.68-1.05) | 1.42 (1.01-2.00) | 1.56 (0.99-2.46) | 0.014 | 1.26 (1.05-1.51) |
| High-risk adenoma | 1 (ref) | 0.88 (0.76-1.01) | 1.21 (0.96-1.53) | 1.14 (0.83-1.58) | 0.188 | 1.09 (0.96-1.24) |
| **Model 3 + glycemic load**^b^ | | | | | | |
| Total adenoma | 1 (ref) | 0.90 (0.82-0.98) | 1.14 (0.98-1.32) | 1.12 (0.91-1.37) | 0.119 | 1.07 (0.98-1.16) |
| Proximal adenoma | 1 (ref) | 0.89 (0.79-1.01) | 1.21 (0.99-1.48) | 1.21 (0.91-1.61) | 0.071 | 1.11 (0.99-1.25) |
| Rectal adenoma | 1 (ref) | 0.83 (0.66-1.03) | 1.36 (0.96-1.93) | 1.39 (0.87-2.23) | 0.056 | 1.20 (1.00-1.46) |
| High-risk adenoma | 1 (ref) | 0.86 (0.74-0.99) | 1.16 (0.91-1.46) | 1.04 (0.75-1.45) | 0.469 | 1.05 (0.92-1.20) |
| **Model 3 + fructose from SSBs^c^** | | | | | | |
| Total adenoma | 1 (ref) | 0.93 (0.81-1.07) | 1.13 (0.90-1.41) | 1.05 (0.77-1.42) | 0.659 | 1.03 (0.91-1.16) |
| Proximal adenoma | 1 (ref) | 0.90 (0.75-1.09) | 1.07 (0.79-1.45) | 0.91 (0.60-1.38) | 0.727 | 0.97 (0.82-1.15) |
| Rectal adenoma | 1 (ref) | 0.80 (0.55-1.16) | 1.51 (0.85-2.66) | 1.71 (0.90-3.26) | 0.014 | 1.33 (1.06-1.66) |
| High-risk adenoma | 1 (ref) | 0.91 (0.73-1.13) | 1.08 (0.76-1.54) | 0.87 (0.53-1.41) | 0.600 | 0.95 (0.78-1.15) |

**Supplementary Table 5. (cont.)**

|  | **Total fructose intake during adolescence, % of calorie** | | | | | | **Per 5% of calorie/d increase** |
| --- | --- | --- | --- | --- | --- | --- | --- |
|  | **Q1** | **Q2** | **Q3** | **Q4** | **Q5** | ***P*_trend_** |  |
| **Model 3^a^** |  |  |  |  |  |  |  |
| Total adenoma | 1 (ref) | 1.05 (0.93-1.19) | 1.14 (1.00-1.30) | 1.19 (1.04-1.36) | 1.20 (1.04-1.39) | 0.006 | 1.17 (1.05-1.31) |
| Distal adenoma | 1 (ref) | 0.97 (0.80-1.18) | 1.10 (0.90-1.35) | 1.21 (0.98-1.48) | 1.25 (1.00-1.56) | 0.014 | 1.24 (1.05-1.47) |
| Rectal adenoma | 1 (ref) | 1.39 (1.01-1.89) | 1.24 (0.90-1.73) | 1.61 (1.15-2.25) | 1.62 (1.14-2.31) | 0.008 | 1.43 (1.10-1.86) |
| High-risk adenoma | 1 (ref) | 1.20 (0.98-1.48) | 1.27 (1.02-1.57) | 1.53 (1.23-1.92) | 1.44 (1.14-1.83) | 0.001 | 1.36 (1.14-1.62) |
| **Model 3 + red meat intake**^b^ | | | | | | | |
| Total adenoma | 1 (ref) | 1.05 (0.93-1.19) | 1.14 (1.00-1.30) | 1.19 (1.03-1.36) | 1.20 (1.03-1.39) | 0.009 | 1.17 (1.04-1.31) |
| Distal adenoma | 1 (ref) | 0.96 (0.79-1.17) | 1.09 (0.89-1.34) | 1.19 (0.96-1.47) | 1.22 (0.96-1.54) | 0.035 | 1.22 (1.01-1.46) |
| Rectal adenoma | 1 (ref) | 1.40 (1.02-1.91) | 1.26 (0.91-1.75) | 1.64 (1.17-2.30) | 1.67 (1.17-2.38) | 0.005 | 1.46 (1.12-1.91) |
| High-risk adenoma | 1 (ref) | 1.21 (0.98-1.49) | 1.28 (1.02-1.59) | 1.55 (1.24-1.94) | 1.47 (1.15-1.88) | 0.001 | 1.38 (1.14-1.66) |
| **Model 3 + prudent dietary pattern**^b^ **(in place of western dietary pattern)** | | | | | | | |
| Total adenoma | 1 (ref) | 1.05 (0.93-1.19) | 1.14 (1.00-1.29) | 1.18 (1.03-1.35) | 1.19 (1.03-1.37) | 0.007 | 1.16 (1.04-1.30) |
| Distal adenoma | 1 (ref) | 0.97 (0.80-1.18) | 1.10 (0.90-1.35) | 1.20 (0.98-1.47) | 1.24 (0.99-1.54) | 0.016 | 1.23 (1.04-1.46) |
| Rectal adenoma | 1 (ref) | 1.38 (1.01-1.88) | 1.22 (0.88-1.69) | 1.56 (1.12-2.18) | 1.56 (1.10-2.20) | 0.014 | 1.38 (1.07-1.79) |
| High-risk adenoma | 1 (ref) | 1.20 (0.97-1.48) | 1.26 (1.01-1.56) | 1.52 (1.22-1.89) | 1.42 (1.12-1.79) | 0.001 | 1.34 (1.12-1.59) |
| **Model 3 + prudent dietary pattern**^b^ **(in addition to western dietary pattern)** | | | | | | | |
| Total adenoma | 1 (ref) | 1.05 (0.93-1.19) | 1.14 (1.00-1.30) | 1.19 (1.04-1.36) | 1.20 (1.04-1.39) | 0.006 | 1.17 (1.05-1.31) |
| Distal adenoma | 1 (ref) | 0.97 (0.80-1.18) | 1.10 (0.90-1.35) | 1.20 (0.98-1.48) | 1.24 (1.00-1.55) | 0.015 | 1.24 (1.04-1.47) |
| Rectal adenoma | 1 (ref) | 1.39 (1.02-1.90) | 1.25 (0.90-1.73) | 1.61 (1.15-2.25) | 1.62 (1.14-2.31) | 0.008 | 1.43 (1.10-1.86) |
| High-risk adenoma | 1 (ref) | 1.21 (0.98-1.49) | 1.27 (1.03-1.58) | 1.55 (1.24-1.94) | 1.46 (1.15-1.85) | 0.001 | 1.37 (1.14-1.63) |
| **Model 3 + SSB intake**^d^ | | | | | | | |
| Total adenoma | 1 (ref) | 1.07 (0.94-1.21) | 1.16 (1.01-1.32) | 1.20 (1.04-1.39) | 1.17 (0.99-1.38) | 0.029 | 1.15 (1.01-1.31) |
| Distal adenoma | 1 (ref) | 0.99 (0.81-1.20) | 1.13 (0.92-1.39) | 1.24 (1.00-1.54) | 1.28 (0.99-1.65) | 0.019 | 1.27 (1.04-1.55) |
| Rectal adenoma | 1 (ref) | 1.41 (1.03-1.93) | 1.26 (0.90-1.77) | 1.61 (1.13-2.28) | 1.46 (0.97-2.21) | 0.069 | 1.33 (0.98-1.80) |
| High-risk adenoma | 1 (ref) | 1.23 (1.00-1.52) | 1.31 (1.05-1.64) | 1.59 (1.27-2.01) | 1.46 (1.11-1.92) | 0.002 | 1.38 (1.12-1.70) |
| **Model 3 + fruit juice intake**^d^ | | | | | | | |
| Total adenoma | 1 (ref) | 1.05 (0.93-1.19) | 1.14 (1.00-1.30) | 1.18 (1.03-1.36) | 1.19 (1.03-1.38) | 0.009 | 1.16 (1.04-1.30) |
| Distal adenoma | 1 (ref) | 0.97 (0.79-1.18) | 1.10 (0.89-1.35) | 1.19 (0.96-1.47) | 1.22 (0.97-1.53) | 0.029 | 1.22 (1.02-1.45) |
| Rectal adenoma | 1 (ref) | 1.42 (1.04-1.95) | 1.29 (0.93-1.79) | 1.67 (1.19-2.34) | 1.68 (1.18-2.39) | 0.005 | 1.46 (1.12-1.90) |
| High-risk adenoma | 1 (ref) | 1.20 (0.97-1.48) | 1.26 (1.01-1.56) | 1.51 (1.21-1.90) | 1.42 (1.11-1.81) | 0.002 | 1.33 (1.11-1.60) |
| **Model 3 + glycemic index**^b^ | | | | | | | |
| Total adenoma | 1 (ref) | 1.05 (0.93-1.20) | 1.14 (1.00-1.30) | 1.19 (1.04-1.36) | 1.19 (1.03-1.37) | 0.009 | 1.16 (1.04-1.29) |
| Distal adenoma | 1 (ref) | 0.97 (0.80-1.18) | 1.10 (0.90-1.35) | 1.21 (0.98-1.48) | 1.24 (0.99-1.55) | 0.017 | 1.23 (1.04-1.47) |
| Rectal adenoma | 1 (ref) | 1.39 (1.02-1.90) | 1.24 (0.90-1.72) | 1.61 (1.15-2.25) | 1.61 (1.13-2.28) | 0.008 | 1.42 (1.10-1.84) |
| High-risk adenoma | 1 (ref) | 1.21 (0.98-1.49) | 1.27 (1.02-1.57) | 1.53 (1.23-1.91) | 1.43 (1.13-1.81) | 0.001 | 1.35 (1.13-1.61) |
| **Model 3 + glycemic load**^b^ | | | | | | | |
| Total adenoma | 1 (ref) | 1.02 (0.89-1.16) | 1.08 (0.94-1.24) | 1.10 (0.95-1.28) | 1.09 (0.92-1.30) | 0.253 | 1.08 (0.94-1.25) |
| Distal adenoma | 1 (ref) | 0.94 (0.77-1.15) | 1.05 (0.84-1.30) | 1.13 (0.89-1.43) | 1.17 (0.89-1.54) | 0.140 | 1.18 (0.95-1.46) |
| Rectal adenoma | 1 (ref) | 1.40 (1.02-1.93) | 1.24 (0.88-1.75) | 1.56 (1.09-2.24) | 1.43 (0.95-2.17) | 0.122 | 1.29 (0.94-1.77) |
| High-risk adenoma | 1 (ref) | 1.16 (0.94-1.44) | 1.19 (0.95-1.50) | 1.42 (1.12-1.81) | 1.28 (0.97-1.69) | 0.051 | 1.24 (1.00-1.53) |

Q, quintile

^a^Adjusted for age, time period of endoscopy, number of endoscopies during the study period (continuous), time since most recent endoscopy (continuous), and reason for current endoscopy, family history of colorectal cancer, menopausal status/menopausal hormone use, current aspirin use ≥2 times/wk, history of type 2 diabetes, adult height (continuous), BMI at age 18 y, current BMI, smoking status at 19 y, current smoking status, alcohol intake at 18–22 y, current alcohol intake, physical activity during grades 9–12, current physical activity, adolescent and current dietary intake (total calorie, total calcium, vitamin D, total folate, total fiber, fruit, vegetables, and dairy), current total red meat intake, a western dietary pattern score during adolescence, and current sugar-sweetened beverage or total fructose intake

^b^As quintiles

^c^Fructose from SSBs: as quintiles for rectal adenoma and categories (<0.6, 0.65-<3.0, ≥3.0% of calories/d) for other adenoma

^d^As categories (<1 serving/wk, 1-6 serving/wk, 1 serving/d, ≥2 serving/d)

**Supplementary Table 6.** Multivariable odds ratios and 95% confidence intervals of CRC precursor according to calorie-adjusted intake of sugars during adolescence in the Nurses’ Health Study II, 1998-2015

|  | **Total fructose intake during adolescence (g/d)** | | | | | | **Per 25 g/d**  **increase** |
| --- | --- | --- | --- | --- | --- | --- | --- |
|  | **Q1** | **Q2** | **Q3** | **Q4** | **Q5** | ***P*_trend_** |  |
| **Total adenoma** |  |  |  |  |  |  |  |
| *N*_cases_/*N*_controls_ | 584/9965 | 544/10005 | 623/9924 | 588/9962 | 570/9978 |  |  |
| Multivariable^a^ | 1 (ref) | 0.98 (0.86-1.11) | 1.18 (1.04-1.34) | 1.15 (1.01-1.32) | 1.17 (1.01-1.35) | 0.008 | 1.11 (1.03-1.21) |
| Proximal adenoma |  |  |  |  |  |  |  |
| *N*_cases_/*N*_controls_ | 319/9784 | 308/9839 | 332/9724 | 302/9779 | 287/9826 |  |  |
| Multivariable | 1 (ref) | 1.02 (0.86-1.20) | 1.16 (0.98-1.38) | 1.10 (0.91-1.32) | 1.09 (0.89-1.32) | 0.340 | 1.05 (0.95-1.18) |
| Distal adenoma |  |  |  |  |  |  |  |
| *N*_cases_/*N*_controls_ | 242/9784 | 212/9839 | 246/9724 | 254/9779 | 251/9826 |  |  |
| Multivariable | 1 (ref) | 0.91 (0.75-1.11) | 1.12 (0.92-1.36) | 1.20 (0.97-1.47) | 1.23 (0.99-1.53) | 0.011 | 1.17 (1.04-1.33) |
| Rectal adenoma |  |  |  |  |  |  |  |
| *N*_cases_/*N*_controls_ | 83/9784 | 78/9839 | 102/9724 | 93/9779 | 102/9826 |  |  |
| Multivariable | 1 (ref) | 1.01 (0.73-1.40) | 1.41 (1.03-1.93) | 1.34 (0.96-1.88) | 1.56 (1.11-2.18) | 0.004 | 1.32 (1.09-1.59) |
| Low-risk adenoma^b^ |  |  |  |  |  |  |  |
| *N*_cases_/*N*_controls_ | 306/9784 | 257/9839 | 294/9724 | 272/9779 | 255/9826 |  |  |
| Multivariable | 1 (ref) | 0.86 (0.72-1.03) | 1.04 (0.87-1.24) | 0.99 (0.81-1.20) | 0.97 (0.79-1.19) | 0.870 | 1.01 (0.90-1.14) |
| High-risk adenoma^b^ |  |  |  |  |  |  |  |
| *N*_cases_/*N*_controls_ | 199/9784 | 204/9839 | 226/9724 | 221/9779 | 232/9826 |  |  |
| Multivariable | 1 (ref) | 1.09 (0.89-1.34) | 1.28 (1.04-1.58) | 1.30 (1.04-1.62) | 1.41 (1.12-1.78) | 0.002 | 1.23 (1.08-1.40) |
| **Total serrated lesions**^c^ |  |  |  |  |  |  |  |
| *N*_cases_/*N*_controls_ | 471/9958 | 470/9965 | 465/9961 | 491/9933 | 458/9989 |  |  |
| Multivariable | 1 (ref) | 1.02 (0.89-1.17) | 1.01 (0.88-1.17) | 1.10 (0.95-1.27) | 1.05 (0.90-1.23) | 0.405 | 1.04 (0.95-1.13) |
| Small serrated lesion^d^ |  |  |  |  |  |  |  |
| *N*_cases_/*N*_controls_ | 396/9958 | 397/9965 | 407/9961 | 427/9933 | 388/9989 |  |  |
| Multivariable | 1 (ref) | 1.03 (0.88-1.20) | 1.06 (0.91-1.24) | 1.15 (0.98-1.35) | 1.07 (0.90-1.27) | 0.277 | 1.05 (0.96-1.16) |
| Large serrated lesion^d^ |  |  |  |  |  |  |  |
| *N*_cases_/*N*_controls_ | 45/9958 | 45/9965 | 31/9961 | 39/9933 | 36/9989 |  |  |
| Multivariable | 1 (ref) | 1.01 (0.66-1.53) | 0.67 (0.41-1.10) | 0.82 (0.51-1.31) | 0.77 (0.48-1.25) | 0.240 | 0.85 (0.64-1.12) |
| **Subtype of CRC precursor** |  |  |  |  |  |  |  |
| Adenoma only |  |  |  |  |  |  |  |
| *N*_cases_/*N*_controls_ | 486/9592 | 451/9628 | 513/9569 | 462/9597 | 477/9613 |  |  |
| Multivariable | 1 (ref) | 0.98 (0.85-1.12) | 1.16 (1.01-1.34) | 1.09 (0.94-1.26) | 1.17 (1.00-1.36) | 0.026 | 1.10 (1.01-1.21) |
| Serrated lesion only |  |  |  |  |  |  |  |
| *N*_cases_/*N*_controls_ | 373/9592 | 377/9628 | 355/9569 | 365/9597 | 365/9613 |  |  |
| Multivariable | 1 (ref) | 1.03 (0.88-1.20) | 0.97 (0.82-1.13) | 1.01 (0.86-1.19) | 1.03 (0.87-1.22) | 0.780 | 1.01 (0.92-1.12) |
| Both adenoma and serrated lesion |  |  |  |  |  |  |  |
| *N*_cases_/*N*_controls_ | 98/9592 | 93/9628 | 110/9569 | 126/9597 | 93/9613 |  |  |
| Multivariable | 1 (ref) | 1.00 (0.74-1.34) | 1.24 (0.92-1.67) | 1.48 (1.09-2.00) | 1.18 (0.84-1.64) | 0.097 | 1.16 (0.97-1.39) |

**Supplementary Table 6. (cont.)**

|  | **Glucose intake during adolescence (g/d)** | | | | | | **Per 25 g/d**  **increase** |
| --- | --- | --- | --- | --- | --- | --- | --- |
|  | **Q1** | **Q2** | **Q3** | **Q4** | **Q5** | ***P*_trend_** |  |
| **Total adenoma** |  |  |  |  |  |  |  |
| *N*_cases_/*N*_controls_ | 586/9962 | 579/9970 | 579/9970 | 593/9956 | 572/9976 |  |  |
| Multivariable^a^ | 1 (ref) | 1.04 (0.92-1.18) | 1.07 (0.94-1.21) | 1.13 (0.99-1.29) | 1.13 (0.98-1.29) | 0.055 | 1.08 (1.00-1.17) |
| Proximal adenoma |  |  |  |  |  |  |  |
| *N*_cases_/*N*_controls_ | 322/9777 | 316/9803 | 316/9780 | 310/9762 | 284/9830 |  |  |
| Multivariable | 1 (ref) | 1.04 (0.88-1.23) | 1.08 (0.91-1.28) | 1.10 (0.92-1.31) | 1.04 (0.86-1.26) | 0.623 | 1.03 (0.92-1.15) |
| Distal adenoma |  |  |  |  |  |  |  |
| *N*_cases_/*N*_controls_ | 238/9777 | 233/9803 | 226/9780 | 253/9762 | 255/9830 |  |  |
| Multivariable | 1 (ref) | 1.03 (0.86-1.25) | 1.02 (0.84-1.25) | 1.19 (0.97-1.45) | 1.22 (0.99-1.51) | 0.028 | 1.15 (1.01-1.30) |
| Rectal adenoma |  |  |  |  |  |  |  |
| *N*_cases_/*N*_controls_ | 84/9777 | 86/9803 | 96/9780 | 84/9762 | 108/9830 |  |  |
| Multivariable | 1 (ref) | 1.07 (0.78-1.47) | 1.23 (0.90-1.68) | 1.11 (0.80-1.55) | 1.51 (1.08-2.11) | 0.016 | 1.27 (1.04-1.55) |
| Low-risk adenoma^b^ |  |  |  |  |  |  |  |
| *N*_cases_/*N*_controls_ | 307/9777 | 267/9803 | 287/9780 | 275/9762 | 248/9830 |  |  |
| Multivariable | 1 (ref) | 0.90 (0.75-1.06) | 1.00 (0.84-1.19) | 0.98 (0.81-1.18) | 0.91 (0.75-1.11) | 0.585 | 0.97 (0.86-1.09) |
| High-risk adenoma^b^ |  |  |  |  |  |  |  |
| *N*_cases_/*N*_controls_ | 194/9777 | 226/9803 | 198/9780 | 225/9762 | 239/9830 |  |  |
| Multivariable | 1 (ref) | 1.23 (1.00-1.51) | 1.11 (0.90-1.38) | 1.31 (1.05-1.63) | 1.43 (1.14-1.79) | 0.002 | 1.22 (1.07-1.39) |
| **Total serrated lesions**^c^ |  |  |  |  |  |  |  |
| *N*_cases_/*N*_controls_ | 455/9982 | 467/9962 | 469/9958 | 494/9935 | 470/9969 |  |  |
| Multivariable | 1 (ref) | 1.06 (0.92-1.22) | 1.07 (0.92-1.23) | 1.14 (0.99-1.33) | 1.12 (0.96-1.31) | 0.110 | 1.08 (0.98-1.18) |
| Small serrated lesion^d^ |  |  |  |  |  |  |  |
| *N*_cases_/*N*_controls_ | 386/9982 | 397/9962 | 405/9958 | 427/9935 | 400/9969 |  |  |
| Multivariable | 1 (ref) | 1.06 (0.91-1.23) | 1.09 (0.93-1.27) | 1.17 (1.00-1.37) | 1.13 (0.95-1.34) | 0.097 | 1.09 (0.99-1.20) |
| Large serrated lesion^d^ |  |  |  |  |  |  |  |
| *N*_cases_/*N*_controls_ | 43/9982 | 41/9962 | 35/9958 | 42/9935 | 35/9969 |  |  |
| Multivariable | 1 (ref) | 0.96 (0.62-1.50) | 0.81 (0.50-1.31) | 0.97 (0.61-1.54) | 0.81 (0.50-1.31) | 0.437 | 0.90 (0.68-1.18) |
| **Subtype of CRC precursor** |  |  |  |  |  |  |  |
| Adenoma only |  |  |  |  |  |  |  |
| *N*_cases_/*N*_controls_ | 495/9598 | 479/9603 | 471/9609 | 473/9582 | 471/9607 |  |  |
| Multivariable | 1 (ref) | 1.02 (0.89-1.16) | 1.03 (0.90-1.18) | 1.06 (0.92-1.23) | 1.09 (0.93-1.27) | 0.230 | 1.06 (0.97-1.15) |
| Serrated lesion only |  |  |  |  |  |  |  |
| *N*_cases_/*N*_controls_ | 364/9598 | 367/9603 | 361/9609 | 374/9582 | 369/9607 |  |  |
| Multivariable | 1 (ref) | 1.03 (0.88-1.20) | 1.01 (0.86-1.18) | 1.06 (0.90-1.25) | 1.07 (0.90-1.27) | 0.401 | 1.04 (0.94-1.15) |
| Both adenoma and serrated lesion |  |  |  |  |  |  |  |
| *N*_cases_/*N*_controls_ | 91/9598 | 100/9603 | 108/9609 | 120/9582 | 101/9607 |  |  |
| Multivariable | 1 (ref) | 1.16 (0.87-1.55) | 1.29 (0.96-1.74) | 1.50 (1.11-2.02) | 1.37 (0.99-1.88) | 0.026 | 1.23 (1.02-1.47) |

**Supplementary Table 6. (cont.)**

|  | **Added sugar intake during adolescence (g/d)** | | | | | | **Per 25 g/d**  **increase** |
| --- | --- | --- | --- | --- | --- | --- | --- |
|  | **Q1** | **Q2** | **Q3** | **Q4** | **Q5** | ***P*_trend_** |  |
| **Total adenoma** |  |  |  |  |  |  |  |
| *N*_cases_/*N*_controls_ | 577/10004 | 582/9939 | 534/10021 | 580/9963 | 636/9907 |  |  |
| Multivariable^a^ | 1 (ref) | 1.02 (0.90-1.15) | 0.93 (0.82-1.05) | 1.03 (0.91-1.17) | 1.17 (1.02-1.34) | 0.015 | 1.05 (1.01-1.10) |
| Proximal adenoma |  |  |  |  |  |  |  |
| *N*_cases_/*N*_controls_ | 301/9815 | 329/9755 | 289/9867 | 295/9771 | 334/9744 |  |  |
| Multivariable | 1 (ref) | 1.10 (0.93-1.30) | 0.97 (0.81-1.15) | 1.01 (0.85-1.21) | 1.18 (0.98-1.42) | 0.162 | 1.04 (0.98-1.10) |
| Distal adenoma |  |  |  |  |  |  |  |
| *N*_cases_/*N*_controls_ | 248/9815 | 229/9755 | 219/9867 | 234/9771 | 275/9744 |  |  |
| Multivariable | 1 (ref) | 0.94 (0.78-1.13) | 0.88 (0.72-1.06) | 0.97 (0.79-1.18) | 1.19 (0.97-1.46) | 0.057 | 1.07 (1.00-1.14) |
| Rectal adenoma |  |  |  |  |  |  |  |
| *N*_cases_/*N*_controls_ | 84/9815 | 86/9755 | 71/9867 | 108/9771 | 109/9744 |  |  |
| Multivariable | 1 (ref) | 1.06 (0.77-1.45) | 0.85 (0.61-1.19) | 1.35 (0.98-1.85) | 1.40 (0.99-1.96) | 0.016 | 1.14 (1.02-1.26) |
| Low-risk adenoma^b^ |  |  |  |  |  |  |  |
| *N*_cases_/*N*_controls_ | 279/9815 | 284/9755 | 257/9867 | 277/9771 | 287/9744 |  |  |
| Multivariable | 1 (ref) | 1.01 (0.85-1.21) | 0.91 (0.75-1.09) | 1.02 (0.85-1.22) | 1.09 (0.89-1.32) | 0.374 | 1.03 (0.97-1.09) |
| High-risk adenoma^b^ |  |  |  |  |  |  |  |
| *N*_cases_/*N*_controls_ | 209/9815 | 211/9755 | 192/9867 | 218/9771 | 252/9744 |  |  |
| Multivariable | 1 (ref) | 1.01 (0.83-1.23) | 0.90 (0.73-1.11) | 1.04 (0.84-1.29) | 1.23 (0.98-1.53) | 0.047 | 1.07 (1.00-1.15) |
| **Total serrated lesions**^c^ |  |  |  |  |  |  |  |
| *N*_cases_/*N*_controls_ | 444/10033 | 453/9952 | 461/9961 | 495/9937 | 502/9923 |  |  |
| Multivariable | 1 (ref) | 1.01 (0.88-1.16) | 1.02 (0.89-1.18) | 1.10 (0.95-1.27) | 1.15 (0.99-1.34) | 0.036 | 1.05 (1.00-1.10) |
| Small serrated lesion^d^ |  |  |  |  |  |  |  |
| *N*_cases_/*N*_controls_ | 379/10033 | 381/9952 | 400/9961 | 430/9937 | 425/9923 |  |  |
| Multivariable | 1 (ref) | 1.00 (0.86-1.16) | 1.05 (0.90-1.22) | 1.13 (0.97-1.32) | 1.15 (0.98-1.36) | 0.036 | 1.06 (1.00-1.11) |
| Large serrated lesion^d^ |  |  |  |  |  |  |  |
| *N*_cases_/*N*_controls_ | 35/10033 | 44/9952 | 34/9961 | 38/9937 | 45/9923 |  |  |
| Multivariable | 1 (ref) | 1.22 (0.77-1.92) | 0.89 (0.54-1.45) | 0.98 (0.61-1.58) | 1.17 (0.73-1.89) | 0.717 | 1.03 (0.89-1.19) |
| **Subtype of CRC precursor** |  |  |  |  |  |  |  |
| Adenoma only |  |  |  |  |  |  |  |
| *N*_cases_/*N*_controls_ | 486/9651 | 477/9591 | 434/9660 | 475/9573 | 517/9524 |  |  |
| Multivariable | 1 (ref) | 1.00 (0.87-1.14) | 0.90 (0.78-1.03) | 1.01 (0.88-1.17) | 1.13 (0.97-1.31) | 0.077 | 1.04 (1.00-1.09) |
| Serrated lesion only |  |  |  |  |  |  |  |
| *N*_cases_/*N*_controls_ | 353/9651 | 348/9591 | 361/9660 | 390/9573 | 383/9524 |  |  |
| Multivariable | 1 (ref) | 0.98 (0.84-1.14) | 0.99 (0.85-1.16) | 1.09 (0.93-1.28) | 1.09 (0.92-1.30) | 0.152 | 1.04 (0.99-1.09) |
| Both adenoma and serrated lesion |  |  |  |  |  |  |  |
| *N*_cases_/*N*_controls_ | 91/9651 | 105/9591 | 100/9660 | 105/9573 | 119/9524 |  |  |
| Multivariable | 1 (ref) | 1.13 (0.85-1.50) | 1.08 (0.80-1.45) | 1.15 (0.86-1.55) | 1.43 (1.05-1.95) | 0.025 | 1.11 (1.01-1.22) |

**Supplementary Table 6. (cont.)**

|  | **Total sugar intake during adolescence (g/d)** | | | | | | **Per 25 g/d**  **increase** |
| --- | --- | --- | --- | --- | --- | --- | --- |
|  | **Q1** | **Q2** | **Q3** | **Q4** | **Q5** | ***P*_trend_** |  |
| **Total adenoma** |  |  |  |  |  |  |  |
| *N*_cases_/*N*_controls_ | 580/9969 | 561/9988 | 586/9963 | 599/9950 | 583/9964 |  |  |
| Multivariable^a^ | 1 (ref) | 0.99 (0.88-1.12) | 1.06 (0.94-1.20) | 1.11 (0.98-1.26) | 1.13 (0.99-1.29) | 0.024 | 1.05 (1.01-1.09) |
| Proximal adenoma |  |  |  |  |  |  |  |
| *N*_cases_/*N*_controls_ | 316/9789 | 303/9799 | 312/9794 | 321/9769 | 296/9801 |  |  |
| Multivariable | 1 (ref) | 0.98 (0.83-1.16) | 1.04 (0.88-1.23) | 1.09 (0.92-1.30) | 1.07 (0.89-1.29) | 0.281 | 1.03 (0.98-1.09) |
| Distal adenoma |  |  |  |  |  |  |  |
| *N*_cases_/*N*_controls_ | 243/9789 | 223/9799 | 231/9794 | 240/9769 | 268/9801 |  |  |
| Multivariable | 1 (ref) | 0.94 (0.78-1.14) | 0.99 (0.81-1.20) | 1.06 (0.87-1.29) | 1.23 (1.00-1.50) | 0.020 | 1.08 (1.01-1.15) |
| Rectal adenoma |  |  |  |  |  |  |  |
| *N*_cases_/*N*_controls_ | 80/9789 | 85/9799 | 92/9794 | 105/9769 | 96/9801 |  |  |
| Multivariable | 1 (ref) | 1.14 (0.83-1.55) | 1.29 (0.95-1.75) | 1.51 (1.11-2.06) | 1.45 (1.06-2.00) | 0.007 | 1.14 (1.04-1.25) |
| Low-risk adenoma^b^ |  |  |  |  |  |  |  |
| *N*_cases_/*N*_controls_ | 283/9789 | 283/9799 | 281/9794 | 287/9769 | 250/9801 |  |  |
| Multivariable | 1 (ref) | 1.02 (0.86-1.21) | 1.02 (0.85-1.22) | 1.07 (0.89-1.28) | 0.98 (0.81-1.20) | 0.976 | 1.00 (0.94-1.06) |
| High-risk adenoma^b^ |  |  |  |  |  |  |  |
| *N*_cases_/*N*_controls_ | 212/9789 | 194/9799 | 207/9794 | 228/9769 | 241/9801 |  |  |
| Multivariable | 1 (ref) | 0.95 (0.78-1.17) | 1.05 (0.86-1.29) | 1.19 (0.97-1.47) | 1.32 (1.07-1.62) | 0.002 | 1.11 (1.04-1.18) |
| **Total serrated lesion**^c^ |  |  |  |  |  |  |  |
| *N*_cases_/*N*_controls_ | 489/9948 | 448/9990 | 489/9927 | 447/9989 | 482/9952 |  |  |
| Multivariable | 1 (ref) | 0.93 (0.81-1.07) | 1.07 (0.93-1.23) | 0.98 (0.85-1.13) | 1.10 (0.95-1.27) | 0.138 | 1.03 (0.99-1.08) |
| Small serrated lesion^d^ |  |  |  |  |  |  |  |
| *N*_cases_/*N*_controls_ | 407/9948 | 382/9990 | 427/9927 | 385/9989 | 414/9952 |  |  |
| Multivariable | 1 (ref) | 0.96 (0.82-1.11) | 1.12 (0.97-1.30) | 1.02 (0.87-1.19) | 1.14 (0.98-1.34) | 0.063 | 1.05 (1.00-1.10) |
| Large serrated lesion^d^ |  |  |  |  |  |  |  |
| *N*_cases_/*N*_controls_ | 47/9948 | 42/9990 | 33/9927 | 43/9989 | 31/9952 |  |  |
| Multivariable | 1 (ref) | 0.91 (0.59-1.39) | 0.74 (0.46-1.18) | 0.94 (0.60-1.46) | 0.70 (0.44-1.13) | 0.198 | 0.91 (0.79-1.05) |
| **Subtype of CRC precursor** |  |  |  |  |  |  |  |
| Adenoma only |  |  |  |  |  |  |  |
| *N*_cases_/*N*_controls_ | 475/9585 | 469/9632 | 473/9587 | 490/9612 | 482/9583 |  |  |
| Multivariable | 1 (ref) | 1.01 (0.88-1.15) | 1.04 (0.90-1.19) | 1.09 (0.94-1.25) | 1.13 (0.97-1.30) | 0.067 | 1.04 (1.00-1.09) |
| Serrated lesion only |  |  |  |  |  |  |  |
| *N*_cases_/*N*_controls_ | 384/9585 | 356/9632 | 376/9587 | 338/9612 | 381/9583 |  |  |
| Multivariable | 1 (ref) | 0.94 (0.80-1.09) | 1.03 (0.88-1.20) | 0.93 (0.79-1.09) | 1.09 (0.93-1.27) | 0.311 | 1.03 (0.98-1.08) |
| Both adenoma and serrated lesion |  |  |  |  |  |  |  |
| *N*_cases_/*N*_controls_ | 105/9585 | 92/9632 | 113/9587 | 109/9612 | 101/9583 |  |  |
| Multivariable | 1 (ref) | 0.91 (0.68-1.21) | 1.19 (0.90-1.57) | 1.20 (0.90-1.61) | 1.18 (0.87-1.60) | 0.115 | 1.08 (0.98-1.18) |

^a^Adjusted for age, time period of endoscopy, number of endoscopies during the study period (continuous), time since most recent endoscopy (continuous), and reason for current endoscopy, family history of colorectal cancer, menopausal status/menopausal hormone use, current aspirin use ≥2 times/wk, history of type 2 diabetes, adult height (continuous), BMI at age 18 y, current BMI, smoking status at 19 y, current smoking status, alcohol intake at 18–22 y, current alcohol intake, physical activity during grades 9–12, current physical activity, adolescent and current dietary intake (total calorie, total calcium, vitamin D, total folate, total fiber, fruit, vegetables, and dairy), current total red meat intake, a western dietary pattern score during adolescence, and current sugar-sweetened beverage or total fructose intake

**Supplementary Table 7.** Multivariable odds ratios and 95% confidence intervals of low- and high-risk colorectal adenoma defined based on alternative definition^a^

|  | **Sugar-sweetened beverage intake during adolescence, servings** | | | | | | **Per 1 serving/d increase** |
| --- | --- | --- | --- | --- | --- | --- | --- |
|  | **<1/week** | **1-6/week** | **1/day** | **≥2/day** |  | ***P*_trend_** |  |
| **Low-risk adenoma^a^** |  |  |  |  |  |  |  |
| *N*_cases_/*N*_controls_ | 701/21206 | 687/21732 | 151/3757 | 89/2257 |  |  |  |
| Multivariable^b^ | 1 (ref) | 0.97 (0.86-1.09) | 1.25 (1.03-1.52) | 1.27 (0.98-1.64) |  | 0.020 | 1.13 (1.02-1.25) |
| **High-risk adenoma**^a^ |  |  |  |  |  |  |  |
| *N*_cases_ | 338 | 305 | 73 | 42 |  |  |  |
| Multivariable | 1 (ref) | 0.85 (0.72-1.01) | 1.18 (0.90-1.55) | 1.07 (0.73-1.57) |  | 0.443 | 1.06 (0.91-1.23) |
| **Proximal** |  |  |  |  |  |  |  |
| *N*_cases_ | 177 | 149 | 39 | 21 |  |  |  |
| Multivariable | 1 (ref) | 0.81 (0.64-1.03) | 1.24 (0.85-1.81) | 1.06 (0.61-1.84) |  | 0.564 | 1.07 (0.86-1.33) |
| **Distal** |  |  |  |  |  |  |  |
| *N*_cases_ | 165 | 163 | 39 | 22 |  |  |  |
| Multivariable | 1 (ref) | 0.90 (0.71-1.15) | 1.20 (0.83-1.74) | 1.04 (0.62-1.75) |  | 0.632 | 1.05 (0.86-1.28) |
| **Rectal** |  |  |  |  |  |  |  |
| *N*_cases_ | 82 | 61 | 21 | 13 |  |  |  |
| Multivariable | 1 (ref) | 0.76 (0.53-1.08) | 1.60 (0.95-2.68) | 1.73 (0.85-3.52) |  | 0.048 | 1.33 (1.00-1.78) |
|  | **Total fructose intake during adolescence, % of calorie** | | | | | | **Per 5% of calorie/d increase** |
|  | **Q1** | **Q2** | **Q3** | **Q4** | **Q5** | ***P*_trend_** |  |
| **Low-risk adenoma** |  |  |  |  |  |  |  |
| *N*_cases_/*N*_controls_ | 346/9795 | 316/9807 | 342/9765 | 307/9776 | 317/9809 |  |  |
| Multivariable | 1 (ref) | 0.95 (0.81-1.12) | 1.07 (0.90-1.26) | 1.00 (0.83-1.20) | 1.07 (0.88-1.30) | 0.410 | 1.07 (0.92-1.24) |
| **High-risk adenoma** |  |  |  |  |  |  |  |
| *N*_cases_ | 131 | 152 | 149 | 168 | 158 |  |  |
| Multivariable | 1 (ref) | 1.24 (0.97-1.59) | 1.25 (0.97-1.62) | 1.49 (1.15-1.94) | 1.40 (1.06-1.86) | 0.012 | 1.30 (1.06-1.60) |
| **Proximal** |  |  |  |  |  |  |  |
| *N*_cases_ | 82 | 72 | 75 | 78 | 79 |  |  |
| Multivariable | 1 (ref) | 0.99 (0.71-1.38) | 1.08 (0.77-1.52) | 1.23 (0.85-1.77) | 1.30 (0.89-1.89) | 0.105 | 1.28 (0.95-1.72) |
| **Distal** |  |  |  |  |  |  |  |
| *N*_cases_ | 62 | 71 | 82 | 85 | 89 |  |  |
| Multivariable | 1 (ref) | 1.17 (0.82-1.68) | 1.37 (0.95-1.98) | 1.46 (1.02-2.10) | 1.44 (0.97-2.14) | 0.052 | 1.33 (1.00-1.78) |
| **Rectal** |  |  |  |  |  |  |  |
| *N*_cases_ | 26 | 39 | 28 | 49 | 35 |  |  |
| Multivariable | 1 (ref) | 1.65 (0.99-2.77) | 1.23 (0.70-2.15) | 2.35 (1.38-4.02) | 1.65 (0.93-2.92) | 0.055 | 1.47 (0.99-2.19) |

^a^Low-risk: small, tubular, and 1 or 2 adenomas, high-risk: large (≥ 1 cm), any villous histology, high-grade dysplasia, or more than 3 adenomas

^b^Adjusted for age, time period of endoscopy, number of endoscopies during the study period (continuous), time since most recent endoscopy (continuous), and reason for current endoscopy, family history of colorectal cancer, menopausal status/menopausal hormone use, current aspirin use ≥2 times/wk, history of type 2 diabetes, adult height (continuous), BMI at age 18 y, current BMI, smoking status at 19 y, current smoking status, alcohol intake at 18–22 y, current alcohol intake, physical activity during grades 9–12, current physical activity, adolescent and current dietary intake (total calorie, total calcium, vitamin D, total folate, total fiber, fruit, vegetables, and dairy), current total red meat intake, a western dietary pattern score during adolescence, and current sugar-sweetened beverage or total fructose intake

**Supplementary Table 8.** Odds ratios and 95% confidence intervals of colorectal adenoma diagnosed by colonoscopy^a^ in the Nurses’ Health Study II, 1998-2015.

|  | **Sugar-sweetened beverage intake during adolescence, servings** | | | | | | **Per 1 serving/d increase** |
| --- | --- | --- | --- | --- | --- | --- | --- |
|  | **<1/week** | **1-6/week** | **1/day** | **≥2/day** |  | ***P*_trend_** |  |
| **Total adenoma** |  |  |  |  |  |  |  |
| *N*_cases_ | 1201/20145 | 1131/20722 | 256/3616 | 153/2189 |  |  |  |
| Multivariable | 1 (ref) | 0.92 (0.84-1.01) | 1.22 (1.05-1.42) | 1.23 (1.00-1.50) |  | 0.008 | 1.11 (1.03-1.21) |
| **Proximal** |  |  |  |  |  |  |  |
| *N*_cases_ | 645/19843 | 591/20442 | 140/3555 | 84/2156 |  |  |  |
| Multivariable | 1 (ref) | 0.90 (0.80-1.02) | 1.26 (1.03-1.55) | 1.29 (0.98-1.70) |  | 0.017 | 1.14 (1.02-1.27) |
| **Distal** |  |  |  |  |  |  |  |
| *N*_cases_ | 491/19843 | 483/20442 | 106/3555 | 59/2156 |  |  |  |
| Multivariable | 1 (ref) | 0.95 (0.83-1.09) | 1.22 (0.97-1.53) | 1.12 (0.82-1.54) |  | 0.241 | 1.08 (0.95-1.22) |
| **Rectal** |  |  |  |  |  |  |  |
| *N*_cases_ | 189/19843 | 162/20442 | 48/3555 | 32/2156 |  |  |  |
| Multivariable | 1 (ref) | 0.83 (0.66-1.04) | 1.49 (1.05-2.12) | 1.70 (1.08-2.69) |  | 0.004 | 1.31 (1.09-1.57) |
| **Low-risk adenoma^b^** |  |  |  |  |  |  |  |
| *N*_cases_ | 571/19843 | 550/20442 | 120/3555 | 67/2156 |  |  |  |
| Multivariable | 1 (ref) | 0.95 (0.83-1.08) | 1.21 (0.98-1.51) | 1.15 (0.86-1.54) |  | 0.157 | 1.09 (0.97-1.22) |
| **High-risk adenoma^b^** |  |  |  |  |  |  |  |
| *N*_cases_ | 448/19843 | 417/20442 | 103/3555 | 60/2156 |  |  |  |
| Multivariable | 1 (ref) | 0.89 (0.77-1.03) | 1.28 (1.01-1.61) | 1.21 (0.88-1.67) |  | 0.082 | 1.12 (0.99-1.27) |
|  | **Total fructose intake during adolescence, % of calorie, quintile** | | | | | | **Per 5% of calorie/d increase** |
|  | **Q1** | **Q2** | **Q3** | **Q4** | **Q5** | ***P*_trend_** |  |
| **Total adenoma** |  |  |  |  |  |  |  |
| *N*_cases_ | 540/9284 | 534/9355 | 562/9307 | 552/9310 | 553/9416 |  |  |
| Multivariable | 1 (ref) | 1.03 (0.90-1.17) | 1.13 (0.99-1.29) | 1.16 (1.01-1.34) | 1.18 (1.02-1.37) | 0.012 | 1.16 (1.03-1.30) |
| **Proximal** |  |  |  |  |  |  |  |
| *N*_cases_ | 298/9146 | 290/9221 | 303/9165 | 288/9176 | 281/9288 |  |  |
| Multivariable | 1 (ref) | 1.02 (0.85-1.21) | 1.12 (0.94-1.34) | 1.11 (0.92-1.35) | 1.10 (0.90-1.36) | 0.267 | 1.09 (0.93-1.28) |
| **Distal** |  |  |  |  |  |  |  |
| *N*_cases_ | 226/9146 | 204/9221 | 230/9165 | 240/9176 | 239/9288 |  |  |
| Multivariable | 1 (ref) | 0.93 (0.76-1.14) | 1.10 (0.89-1.35) | 1.20 (0.97-1.48) | 1.21 (0.97-1.52) | 0.023 | 1.23 (1.03-1.47) |
| **Rectal** |  |  |  |  |  |  |  |
| *N*_cases_ | 73/9146 | 89/9221 | 80/9165 | 93/9176 | 96/9288 |  |  |
| Multivariable | 1 (ref) | 1.32 (0.96-1.83) | 1.25 (0.89-1.75) | 1.58 (1.12-2.24) | 1.68 (1.17-2.42) | 0.004 | 1.49 (1.13-1.96) |
| **Low-risk adenoma** |  |  |  |  |  |  |  |
| *N*_cases_ | 283/9146 | 251/9221 | 278/9165 | 234/9176 | 262/9288 |  |  |
| Multivariable | 1 (ref) | 0.90 (0.75-1.08) | 1.03 (0.86-1.25) | 0.90 (0.73-1.10) | 1.03 (0.83-1.27) | 0.778 | 1.02 (0.86-1.22) |
| **High-risk adenoma** |  |  |  |  |  |  |  |
| *N*_cases_ | 183/9146 | 199/9221 | 206/9165 | 229/9176 | 211/9288 |  |  |
| Multivariable | 1 (ref) | 1.16 (0.93-1.43) | 1.26 (1.01-1.57) | 1.50 (1.20-1.88) | 1.40 (1.10-1.79) | 0.002 | 1.34 (1.11-1.61) |

^a^After excluding participants with sigmoidoscopy only. Data were adjusted for age, time period of endoscopy, number of endoscopies during the study period (continuous), time since most recent endoscopy (continuous), and reason for current endoscopy, family history of colorectal cancer, menopausal status/menopausal hormone use, current aspirin use ≥2 times/wk, history of type 2 diabetes, adult height (continuous), BMI at age 18 y, current BMI, smoking status at 19 y, current smoking status, alcohol intake at 18–22 y, current alcohol intake, physical activity during 9^th^–12^th^ grades, and current physical activity, adolescent and current dietary intake (total calorie, total calcium, vitamin D, total folate, total fiber, fruits, vegetables, and dairy), current total red meat intake, a western dietary pattern score during adolescence, and current sugar-sweetened beverage or total fructose intake

^b^Low-risk: small, tubular, and 1 adenoma, high-risk: large (≥ 1 cm), any villous histology, high-grade dysplasia, or more than 2 adenomas

**Supplementary Table 9.** Multivariable odds ratios (ORs) and 95% confidence intervals (CIs) of total adenoma according to sugar-sweetened beverage and total fructose intake during adolescence by family history and lifestyle factors in the Nurses’ Health Study II, 1998-2015

|  | **Sugar-sweetened beverage intake^a^** | | |  | **Total fructose intake^b^** | | |
| --- | --- | --- | --- | --- | --- | --- | --- |
|  | **N_cases_/N_controls_** | **OR (95% CI)^c^** | ***P*_inter_** |  | **N_cases_/N_controls_** | **OR (95% CI)^c^** | ***P*_inter_** |
| **Family history of CRC** |  |  |  |  |  |  |  |
| No | 284/4378 | 1.20 (1.02-1.41) | 0.51 |  | 380/7344 | 1.10 (0.92-1.31) | 0.21 |
| Yes | 144/1756 | 1.18 (0.94-1.49) |  |  | 200/2624 | 1.42 (1.10-1.85) |  |
| ***Lifestyle factors during adolescence*** |  |  |  |  |  |  |  |
| **BMI** |  |  |  |  |  |  |  |
| ≤22.5 kg/m^2^ | 325/4832 | 1.20 (1.03-1.39) | 0.86 |  | 433/7876 | 1.23 (1.04-1.45) | 0.70 |
| >22.5 kg/m^2^ | 100/1257 | 1.24 (0.95-1.61) |  |  | 143/2011 | 1.19 (0.89-1.60) |  |
| **Physical activity^d^** |  |  |  |  |  |  |  |
| Low | 272/3675 | 1.22 (1.04-1.44) | 0.15 |  | 388/5987 | 1.23 (1.04-1.47) | 0.50 |
| High | 142/2152 | 1.09 (0.86-1.38) |  |  | 171/3454 | 1.10 (0.84-1.45) |  |
| **Smoking** |  |  |  |  |  |  |  |
| No | 289/4324 | 1.16 (0.99-1.35) | 0.31 |  | 433/7620 | 1.20 (1.01-1.42) | 0.61 |
| Yex | 139/1810 | 1.37 (1.06-1.77) |  |  | 147/2348 | 1.22 (0.91-1.64) |  |
| **Alcohol intake** |  |  |  |  |  |  |  |
| <5 g/d | 234/3285 | 1.27 (1.06-1.51) | 0.66 |  | 345/5846 | 1.21 (1.01-1.46) | 0.90 |
| ≥5 g/d | 193/2819 | 1.14 (0.93-1.39) |  |  | 234/4062 | 1.18 (0.94-1.48) |  |
| ***Dietary intake during adolescence*** |  |  |  |  |  |  |  |
| **Fruit intake^e^** |  |  |  |  |  |  |  |
| Low | 277/3683 | 1.33 (1.11-1.59) | 0.024 |  | 253/3503 | 1.51 (1.26-1.82) | <0.001 |
| High | 151/2451 | 1.06 (0.86-1.30) |  |  | 327/6465 | 0.88 (0.70-1.10) |  |
| **Fruit juice intake^e^** |  |  |  |  |  |  |  |
| Low | 226/3278 | 1.15 (0.96-1.39) | 0.98 |  | 224/3737 | 1.21 (1.00-1.48) | 0.75 |
| High | 201/2846 | 1.25 (1.04-1.51) |  |  | 356/6227 | 1.19 (0.93-1.51) |  |
| **Fruit/fruit juice intake^e^** |  |  |  |  |  |  |  |
| High/low | 50/834 | 0.99 (0.69-1.43) | 0.016 |  | 79/1691 | 0.74 (0.52-1.05) | <0.001 |
| High/high | 101/1617 | 1.10 (0.85-1.42) |  |  | 248/4774 | 0.96 (0.67-1.38) |  |
| Low/low | 176/2444 | 1.22 (0.98-1.53) |  |  | 145/2046 | 1.52 (1.20-1.91) |  |
| Low/high | 100/1229 | 1.61 (1.19-2.18) |  |  | 108/1453 | 1.58 (1.12-2.24) |  |
| **Vegetable intake^e^** |  |  |  |  |  |  |  |
| Low | 260/3282 | 1.40 (1.16-1.68) | 0.012 |  | 286/4464 | 1.37 (1.13-1.67) | 0.095 |
| High | 168/2852 | 1.00 (0.82-1.21) |  |  | 294/5504 | 1.00 (0.80-1.23) |  |
| **Fiber intake^e^** |  |  |  |  |  |  |  |
| Low | 324/4060 | 1.39 (1.17-1.64) | 0.008 |  | 274/3927 | 1.37 (1.13-1.65) | 0.029 |
| High | 104/2074 | 0.89 (0.70-1.12) |  |  | 306/6041 | 1.04 (0.82-1.31) |  |
| **Prudent dietary pattern score^e^** |  |  |  |  |  |  |  |
| Low | 306/3976 | 1.31 (1.11-1.55) | 0.11 |  | 304/4456 | 1.37 (1.14-1.65) | 0.012 |
| High | 103/1791 | 1.01 (0.80-1.28) |  |  | 252/5026 | 0.98 (0.79-1.20) |  |
| **Western dietary pattern score^e^** |  |  |  |  |  |  |  |
| Low | 201/2837 | 1.21 (1.00-1.47) | 0.92 |  | 271/5404 | 0.94 (0.76-1.16) | 0.002 |
| High | 208/2930 | 1.19 (0.99-1.43) |  |  | 285/4078 | 1.44 (1.19-1.74) |  |

CRC, colorectal cancer; BMI, body mass index

^a^Comparison of sugar-sweetened bevarage intake between categories of ≥1 serving/d vs. <1 serving/wk (referent)

^b^Comparison of total fructose intake between the highest vs. lowest quintiles (referent)

^c^Adjusted for age, time period of endoscopy, number of endoscopies during the study period (continuous), time since most recent endoscopy (continuous), and reason for current endoscopy, family history of colorectal cancer, menopausal status/menopausal hormone use, current aspirin use ≥2 times/wk, history of type 2 diabetes, adult height (continuous), BMI at age 18 y, current BMI, smoking status at 19 y, current smoking status, alcohol intake at 18–22 y, current alcohol intake, physical activity during 9^th^–12^th^ grades, and current physical activity, adolescent and current dietary intake (total calorie, total calcium, vitamin D, total folate, total fiber, fruits, vegetables, and dairy), current total red meat intake, a western dietary pattern score during adolescence, and current sugar-sweetened beverage or total fructose intake except for the stratifying variable of each stratum

^d^High physical activity was defined as the highest tertile (≥59 MET-hr/wk), and low physical activity as 2 bottom tertiles (<59 MET-hr/wk)

^e^Cut-off values were median intake value (fruits, 1.3 serving/d; fruit juice, 0.4 serving/d; vegetables, 2.8 serving/d)

**Supplementary Table 10.** Multivariable odds ratios (ORs) and 95% confidence intervals (CIs) of high-risk colorectal adenoma^a^ according to sugar-sweetened beverage and total fructose intake during adolescence by family history and lifestyle factors in the Nurses’ Health Study II, 1998-2015

|  | **Sugar-sweetened beverage intake^b^** | | |  | **Total fructose intake^b^** | | |
| --- | --- | --- | --- | --- | --- | --- | --- |
|  | **N_cases_/N_controls_** | **OR (95% CI)^c^** | ***P*_inter_** |  | **N_cases_/N_controls_** | **OR (95% CI)^c^** | ***P*_inter_** |
| **Family history of CRC** |  |  |  |  |  |  |  |
| No | 114/4313 | 1.25 (0.97-1.62) | 0.98 |  | 145/7261 | 1.31 (0.99-1.74) | 0.32 |
| Yes | 56/1701 | 1.19 (0.82-1.72) |  |  | 77/2548 | 1.75 (1.13-2.72) |  |
| ***Lifestyle factors during adolescence*** |  |  |  |  |  |  |  |
| **BMI** |  |  |  |  |  |  |  |
| ≤22.5 kg/m^2^ | 129/4743 | 1.17 (0.92-1.50) | 0.61 |  | 161/7768 | 1.39 (1.06-1.82) | 0.43 |
| >22.5 kg/m^2^ | 39/1227 | 1.54 (1.00-2.36) |  |  | 60/1962 | 1.65 (0.99-2.74) |  |
| **Physical activity^d^** |  |  |  |  |  |  |  |
| Low | 104/3591 | 1.21 (0.93-1.57) | 0.91 |  | 151/5882 | 1.52 (1.14-2.03) | 0.48 |
| High | 59/2116 | 1.19 (0.81-1.74) |  |  | 64/3400 | 1.25 (0.81-1.94) |  |
| **Alcohol intake** |  |  |  |  |  |  |  |
| <5 g/d | 95/3222 | 1.32 (1.00-1.74) | 0.63 |  | 138/5743 | 1.41 (1.04-1.91) | 0.32 |
| ≥5 g/d | 75/2763 | 1.16 (0.84-1.60) |  |  | 83/4006 | 1.43 (0.97-2.11) |  |
| ***Dietary intake during adolescence*** |  |  |  |  |  |  |  |
| **Fruit intake^e^** |  |  |  |  |  |  |  |
| Low | 112/3611 | 1.36 (1.03-1.80) | 0.10 |  | 102/3437 | 1.77 (1.31-2.40) | 0.057 |
| High | 58/2403 | 1.09 (0.79-1.52) |  |  | 120/6372 | 1.04 (0.70-1.53) |  |
| **Fruit juice intake^e^** |  |  |  |  |  |  |  |
| Low | 91/3225 | 1.30 (0.97-1.75) | 0.66 |  | 82/3681 | 1.46 (1.06-2.02) | 0.25 |
| High | 78/2779 | 1.18 (0.87-1.59) |  |  | 140/6124 | 1.46 (0.97-2.21) |  |
| **Vegetable intake^e^** |  |  |  |  |  |  |  |
| Low | 115/3215 | 1.74 (1.31-2.31) | 0.005 |  | 113/4405 | 1.81 (1.31-2.50) | 0.047 |
| High | 55/2799 | 0.76 (0.55-1.06) |  |  | 109/5404 | 1.08 (0.76-1.53) |  |
| **Fiber intake^e^** |  |  |  |  |  |  |  |
| Low | 135/3971 | 1.47 (1.12-1.93) | 0.008 |  | 105/3852 | 1.56 (1.14-2.14) | 0.11 |
| High | 35/2043 | 0.80 (0.55-1.18) |  |  | 117/5957 | 1.29 (0.88-1.89) |  |
| **Prudent dietary pattern score^e^** |  |  |  |  |  |  |  |
| Low | 126/3892 | 1.44 (1.11-1.87) | 0.13 |  | 124/4382 | 1.70 (1.26-2.31) | 0.044 |
| High | 38/1760 | 0.97 (0.67-1.42) |  |  | 92/4949 | 1.12 (0.79-1.58) |  |
| **Western dietary pattern score^e^** |  |  |  |  |  |  |  |
| Low | 76/2777 | 1.18 (0.86-1.61) | 0.61 |  | 107/5332 | 1.23 (0.86-1.75) | 0.34 |
| High | 88/2875 | 1.34 (1.00-1.79) |  |  | 109/3999 | 1.62 (1.19-2.21) |  |

CRC, colorectal cancer; BMI, body mass index

^a^Comparison of sugar-sweetened bevarage intake between categories of ≥1 serving/d vs. <1 serving/wk (referent)

^b^Comparison of total fructose intake between the highest vs. lowest quintiles (referent)

^c^Adjusted for age, time period of endoscopy, number of endoscopies during the study period (continuous), time since most recent endoscopy (continuous), and reason for current endoscopy, family history of colorectal cancer, menopausal status/menopausal hormone use, current aspirin use ≥2 times/wk, history of type 2 diabetes, adult height (continuous), BMI at age 18 y, current BMI, smoking status at 19 y, current smoking status, alcohol intake at 18–22 y, current alcohol intake, physical activity during 9^th^–12^th^ grades, and current physical activity, adolescent and current dietary intake (total calorie, total calcium, vitamin D, total folate, total fiber, fruits, vegetables, and dairy), current total red meat intake, a western dietary pattern score during adolescence, and current sugar-sweetened beverage or total fructose intake except for the stratifying variable of each stratum

^d^High physical activity was defined as the highest tertile (≥59 MET-hr/wk), and low physical activity as 2 bottom tertiles (<59 MET-hr/wk)

^e^Cut-off values were median intake value (fruits, 1.3 serving/d; fruit juice, 0.4 serving/d; vegetables, 2.8 serving/d)

**Supplementary Table 11.** Odds ratios (ORs) and 95% confidence intervals (CIs) of early- and late-onset colorectal adenoma according to sugar-sweetened beverage and total fructose intake during adolescence in the Nurses’ Health Study II, 1998-2015

|  | **Sugar-sweetened beverage^a^** | | |  | **Total fructose^b^** | | |
| --- | --- | --- | --- | --- | --- | --- | --- |
|  | ***N*_cases_/*N*_controls_** | **Per 1 serving/d increase** | ***P*_inter_** |  | ***N*_cases_/*N*_controls_** | **Per 5% of  calorie/d increase** | ***P*_inter_** |
| **Total adenoma** |  |  |  |  |  |  |  |
| Early-onset (<55 y) |  |  |  |  |  |  |  |
| Overall | 1792 | 1.09 (0.98-1.20) |  |  | 1792 | 1.24 (1.07-1.43) |  |
| Family history^c^, no | 1201/36420 | 1.11 (0.92-1.35) | 0.35 |  | 1201/36420 | 1.17 (0.98-1.39) | 0.50 |
| Family history, yes | 591/13414 | 1.07 (0.82-1.39) |  |  | 591/13414 | 1.35 (1.04-1.74) |  |
| Late-onset (≥55y) |  |  |  |  |  |  |  |
| Overall | 1117 | 1.11 (0.98-1.26) |  |  | 1117 | 1.08 (0.90-1.29) |  |
| Family history, no | 793/36420 | 1.16 (0.92-1.45) | 0.97 |  | 793/36420 | 0.99 (0.80-1.22) | 0.23 |
| Family history, yes | 324/13414 | 1.25 (0.88-1.79) |  |  | 324/13414 | 1.35 (0.96-1.91) |  |
| **High-risk adenoma** |  |  |  |  |  |  |  |
| Early-onset (<55 y) |  |  |  |  |  |  |  |
| Overall | 665 | 1.09 (0.93-1.28) |  |  | 665 | 1.39 (1.10-1.74) |  |
| Family history, no | 444/35916 | 1.20 (0.87-1.64) | 0.81 |  | 444/35916 | 1.29 (0.98-1.70) | 0.36 |
| Family history, yes | 221/13036 | 1.01 (0.66-1.55) |  |  | 221/13036 | 1.55 (1.01-2.37) |  |
| Late-onset (≥55y) |  |  |  |  |  |  |  |
| Overall | 417 | 1.11 (0.91-1.35) |  |  | 417 | 1.32 (0.99-1.74) |  |
| Family history, no | 298/35916 | 1.14 (0.80-1.63) | 0.83 |  | 298/35916 | 1.23 (0.89-1.69) | 0.63 |
| Family history, yes | 119/13036 | 1.32 (0.73-2.36) |  |  | 119/13036 | 1.71 (0.95-3.11) |  |

Data were adjusted for age, time period of endoscopy, number of endoscopies during the study period (continuous), time since most recent endoscopy (continuous), and reason for current endoscopy, family history of colorectal cancer, menopausal status/menopausal hormone use, current aspirin use ≥2 times/wk, history of type 2 diabetes, adult height (continuous), BMI at age 18 y, current BMI, smoking status at 19 y, current smoking status, alcohol intake at 18–22 y, current alcohol intake, physical activity during grades 9–12, current physical activity, adolescent and current dietary intake (total calorie, total calcium, vitamin D, total folate, total fiber, fruit, vegetables, and dairy), current total red meat intake, a western dietary pattern score during adolescence, current sugar-sweetened beverage or total fructose intake, and family history of colorectal cancer (yes/no) in the models for overall participants only

^a^Comparison of sugar-sweetened bevarage intake between categories of ≥1 serving/d and <1 serving/wk (referent)

^b^Comparison of total fructose intake between the highest and lowest quintiles (referent)

^c^Family history of colorectal cancer in first degree relatives

**Supplemtental Table 12.** Odds ratios and 95% confidence intervals of CRC precursor according to joint categories of sugar-sweetened beverage and total fructose intake during adolescence and adulthood in the Nurses’ Health Study II, 1998-2015^a^

|  |  | **Joint categories of intake (adolescence/adulthood)** | | | |
| --- | --- | --- | --- | --- | --- |
|  |  | **Low/low** | **Low/high** | **High/low** | **High/high** |
| **Sugar-sweetened beverage** |  |  |  |  |  |
| Total adenoma | N_cases_/N_controls_ | 2292/40113 | 189/3587 | 330/4606 | 98/1528 |
|  | OR (95% CI)^a^ | 1 (ref) | 0.91 (0.77-1.07) | 1.27 (1.11-1.44) | 1.13 (0.90-1.42) |
| Rectal adenoma | N_cases_/N_controls_ | 343/39422 | 32/3516 | 65/4518 | 18/1496 |
|  | OR (95% CI)^a^ | 1 (ref) | 0.95 (0.63-1.42) | 1.73 (1.30-2.32) | 1.32 (0.78-2.24) |
| High-risk adenoma^b^ | N_cases_/N_controls_ | 837/39422 | 75/3516 | 123/4518 | 47/1496 |
|  | OR (95% CI)^a^ | 1 (ref) | 0.95 (0.73-1.23) | 1.29 (1.05-1.59) | 1.40 (1.00-1.96) |
| **Total fructose** |  |  |  |  |  |
| Total adenoma | N_cases_/N_controls_ | 1196/20365 | 542/9543 | 603/9564 | 568/10362 |
|  | OR (95% CI)^a^ | 1 (ref) | 1.06 (0.94-1.18) | 1.16 (1.04-1.30) | 1.08 (0.96-1.22) |
| Rectal adenoma | N_cases_/N_controls_ | 180/20005 | 78/9362 | 105/9396 | 95/10189 |
|  | OR (95% CI)^a^ | 1 (ref) | 0.91 (0.68-1.20) | 1.34 (1.02-1.75) | 1.10 (0.82-1.48) |
| High-risk adenoma^b^ | N_cases_/N_controls_ | 427/20005 | 191/9362 | 245/9396 | 219/10189 |
|  | OR (95% CI)^a^ | 1 (ref) | 1.03 (0.86-1.24) | 1.34 (1.12-1.60) | 1.18 (0.97-1.43) |

^a^For the category of sugar-sweetened beverage intake ≥1 serving/day with the category of <1 serving/week as referent. Data were adjusted for age, time period of endoscopy, number of endoscopies during the study period (continuous), time since most recent endoscopy (continuous), and reason for current endoscopy, family history of colorectal cancer, menopausal status/menopausal hormone use, current aspirin use ≥2 times/wk, history of type 2 diabetes, adult height (continuous), BMI at age 18 y, current BMI, smoking status at 19 y, current smoking status, alcohol intake at 18–22 y, current alcohol intake, physical activity during grades 9–12, current physical activity, adolescent and current dietary intake (total calorie, total calcium, vitamin D, total folate, total fiber, fruit, vegetables, and dairy), current total red meat intake, a western dietary pattern score during adolescence, and current sugar-sweetened beverage or total fructose intake

^b^Large (≥ 1 cm) or villous adenoma or more than two adenoma

**Supplementary Table 13.** Multivariable odds ratios (ORs) and 95% confidence intervals (CIs) associated with substitution of alternative beverages and foods for sugar-sweetened beverages in the Nurses’ Health Study II, 1998-2015

|  | **Alternative beverage or food for sugar-sweetened beverages (alternative:SSBs)** | | | | | | | |
| --- | --- | --- | --- | --- | --- | --- | --- | --- |
|  | **Fruit juice (1:1)** | |  | **Fruit (2:2)** | |  | **Total dairy (2:2)** | |
|  | OR (95% CI) | P |  | OR (95% CI) | P |  | OR (95% CI) | P |
| Total adenoma | 0.95 (0.82-1.10) | 0.503 |  | 0.86 (0.68-1.09) | 0.219 |  | 0.82 (0.64-1.05) | 0.109 |
| Proximal adenoma | 0.90 (0.73-1.10) | 0.302 |  | 0.75 (0.54-1.05) | 0.093 |  | 0.80 (0.58-1.12) | 0.189 |
| Distal adenoma | 1.03 (0.82-1.29) | 0.800 |  | 1.04 (0.73-1.49) | 0.814 |  | 0.94 (0.65-1.36) | 0.742 |
| Rectal adenoma | 0.81 (0.57-1.16) | 0.252 |  | 0.62 (0.35-1.08) | 0.089 |  | 0.53 (0.29-0.94) | 0.030 |
| Low-risk adenoma | 0.92 (0.74-1.14) | 0.454 |  | 0.94 (0.67-1.32) | 0.731 |  | 0.89 (0.63-1.26) | 0.497 |
| High-risk adenoma | 1.00 (0.79-1.27) | 0.987 |  | 0.83 (0.57-1.22) | 0.339 |  | 0.77 (0.52-1.13) | 0.184 |

Data were adjusted for age, time period of endoscopy, number of endoscopies during the study period (continuous), time since most recent endoscopy (continuous), and reason for current endoscopy, family history of colorectal cancer, menopausal status/menopausal hormone use, current aspirin use ≥2 times/wk, history of type 2 diabetes, adult height (continuous), BMI at age 18 y, current BMI, smoking status at 19 y, current smoking status, alcohol intake at 18–22 y, current alcohol intake, physical activity during grades 9–12, current physical activity, adolescent and current dietary intake (total calorie, total calcium, vitamin D, total folate, total fiber, fruit, vegetables, and dairy), current total red meat intake, a western dietary pattern score during adolescence, and current sugar-sweetened beverage or total fructose intake

**Supplementary Fig 1.** Joint associations of sugar-sweetened beverage and total fructose intake during adolescence and adulthood with colorectal adenoma in the Nurses’ Health Study II, 1998-2015

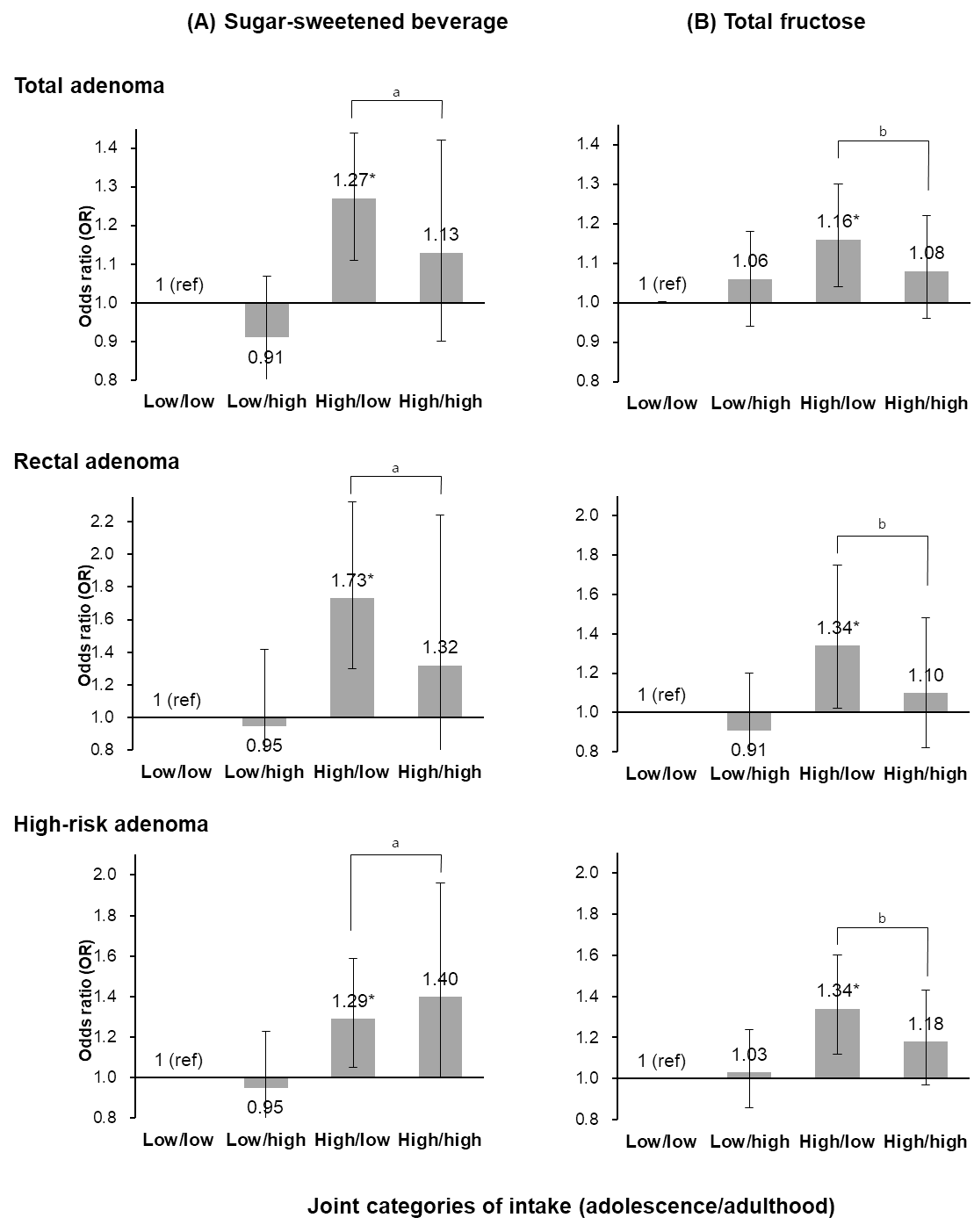

Data were adjusted for age, time period of endoscopy, number of endoscopies during the study period (continuous), time since most recent endoscopy (continuous), and reason for current endoscopy, family history of colorectal cancer, menopausal status/menopausal hormone use, current aspirin use ≥2 times/wk, history of type 2 diabetes, adult height (continuous), BMI at age 18 y, current BMI, smoking status at 19 y, current smoking status, alcohol intake at 18–22 y, current alcohol intake, physical activity during grades 9–12, current physical activity, adolescent and current dietary intake (total calorie, total calcium, vitamin D, total folate, total fiber, fruit, vegetables, and dairy), current total red meat intake, and a western dietary pattern score during adolescence

^*^P ≤ 0.03

^a^P ≥ 0.34

^b^P ≥ 0.19
